# *“I had to learn to trust my body again”:* Exploring the emotional and behavioural impact of wearable activity tracker discontinuation and reasons for removal

**DOI:** 10.64898/2026.05.14.26353189

**Authors:** Gabrielle Humphreys, Sam Jensen, Kieran Manchester, Nilihan Sanal-Hayes, Ashley Gluchowski

## Abstract

While wearable activity trackers (WATs) are widely used in the present day, with device ownership increasing, some individuals subsequently discontinue device use. Existing research primarily examines the initiation and maintenance of device use, with less focus on device discontinuation. Examining this phenomenon can provide valuable insight into human-computer interactions and habit reversal. Therefore, the current study examined the perceived emotional and behavioural impact of WAT discontinuation, alongside reasons for this action in former WAT users.

Fifteen former WAT users (9 female, aged 23 to 56 years) who reported either full or partial device discontinuation were interviewed. Three themes and nine sub-themes were identified which detailed the impacts of device discontinuation. Participants reported a mindset shift around one’s body image, exercise performance and exercise motivation. Device discontinuation removed numerical feedback provision which led to participants gaining bodily intuition and a sense of freedom. However, discontinuation also resulted in short-term negative emotions including frustration around the loss of external praise and envy in current WAT users. Current findings hold important implications around digital safety from user perspective, highlighting the need for guidance around healthy WAT use and vulnerable user profiles. More broadly, findings also raise the need for physical activity promotion whilst protecting individuals’ well-being.

**Author summary:** Wearable activity trackers, such as smartwatches, rings, and chest straps, are growing in popularity, but some individuals choose to discontinue device use. Research often focusses on why people use devices, neglecting why some people stop using them. In the current study, we examined the perceived emotional and behavioural impact of device discontinuance in former wearable activity tracker users. Device discontinuation left participants feeling less pressured to exercise and no longer doing so simply to receive positive data, which resulted in a sense of freedom. Participants reported enjoying exercise more and being kinder to themselves following device discontinuation. Although, they also felt frustrated and sometimes envious over the loss of this feedback, and some participants struggled to initially adapt to life without their device. Our findings showed the need for guidance and support provision over healthy use of wearable activity trackers.

## 1. Introduction

Wearable activity trackers (WATs) are electronic devices worn on the body which allow users continuous self-tracking of health data [1]. The most common WATs are smartwatches, with an estimated 454 million users worldwide [2], although WATs may also be rings, chest straps, or wrist straps. Leading WAT brands are Fitbit, Garmin, Misfit, Apple and Polar [3]. Typical data provided by WATs focusses on step count, sleep score, heart rate, and activity performance [4]. WATs can be viewed as health and activity promotion tools, with Ferguson et al. [5] identifying an average increase of 1,800 steps and 40 minutes of walking per day in WAT users across 39 relevant papers, which led to approximately 1kg of weight reduction for users. These behavioural outcomes are hypothesised to occur via increasing a user’s extrinsic motivation via providing encouragement to reach set numerical thresholds [6–8].

WATs are known for their glanceability, with low access time but high access frequency [9]. These quick yet repeated viewings of objective data on users’ activity can result in the quantified self and a loss of bodily intuition [10,11]. This constant provision of feedback can result in device dependency and fear over WAT removal in some [6]. However, this is not always the case with others either temporarily or permanently discontinuing use [12]. Device discontinuation - when an individual stops using a piece of technology - can be categorised over the stages of device use into rejection in the early phase of device use, regressive discontinuance within the adoption phase of a device, and quitting or temporary discontinuance after continued use [13]. While discontinuation of a device is a crucial stage in a behavioural cycle, existing literature tends to focus on the adoption and maintenance of WAT use, with limited research into this [12,14,15].

Most existing literature on WAT discontinuance focusses on reasons for this action. Jeong et al. [16] examined this via content analysis of Craigslist posts selling WATs, identifying reasons including a gap between expectations and actual usage, a change in health status, change in activity, and an accomplishment of a goal. Alongside a change in health status or life circumstance, Epstein et al. [17] reported that discontinuation occurred due to the cost of inputting data, the cost of having data, discomfort experienced from WAT data, concerns on data accuracy, and users feeling like they had learned enough from their WAT. The scope of this study also extended beyond health tracking tools to well-being and financial tools. Furthermore, Esmonde [18] highlighted that some females discontinue WAT use in running as a ‘resistance to datafication’. Device removal was due to feeling that data volume was excessive and acknowledging that data receival often led to a re-evaluation of an activity [18].

Examining the impact of running both with and without a WAT in amateur females, Tapia [19] reported that users felt that removing a device felt freeing and led to greater enjoyment of the activity. Users reported that tracking a run led to distraction from the activity, and instead they focused on the metrics provided by a WAT [19]. Although the removal of WATs led to less distraction, it also led to feelings of less reward by removing external praise, highlighting both positive and negative outcomes of WAT discontinuation [19]. Furthermore, removing a WAT might not have immediate impact; Clark, Southerton and Driller [20] reported WAT removal still left lingering gestures of wrist checking, altered routines that formed from WAT data remaining in place, and participants still having a heightened awareness over their bodies. Notably, these participants had only worn their Fitbit devices for 7-10 days for the purpose of this study meaning their effects may be amplified further in long-term users - something the current study aims to investigate. This continued use of knowledge following WAT removal was also reported by Epstein et al. [17], for example, with a participant feeling ‘data had done its job’ as they could now comfortably run 5 kilometres. However, the impact of WAT removal remained unclear here, with conflicting outcomes reported of guilt around habit formation, frustration around WATs, freedom in activity and no major emotional effects all being reported [17]. Given the ever-growing popularity of WAT use in the present day, those who have discontinued WAT use report being ‘a significant minority’ [21]. This suggests that stopping use of a WAT is going against normalised and expected behaviour in society. Therefore, examining why some individuals do remove their devices and resist this social norm in the present day is particularly important.

The current study aimed to examine the perceived emotional and behavioural impacts of device discontinuation, alongside reasons for discontinuance. This is the first qualitative study to our knowledge which examined both reasons for and perceived impacts of removal together.

Furthermore, this fills a crucial gap identified in the literature [12,14,15,21] with more research needed in device discontinuation as adoption and maintenance remain prioritised. Most existing literature focusses on smartwatch use in runners [17,19]. Therefore, to further extend the novelty of this study, all types of WATs – including chest straps and smart rings alongside smartwatches – were eligible for the current study. Alongside this, all types of previous WAT use – for different types of physical activity or no physical activity tracking at all – were welcomed. This broadening of devices and behaviours will provide greater understanding of the quantified self and choosing to reject the provision of this information in such a data driven society.

## 2. Methods

### 2.1 Study design

Building on the work of Clark, Southerton and Driller [20] and the uncertainty expressed around device removal of full-time WAT uses [6], the current study adopts a qualitative, exploratory design using semi-structured interviews to examine the perceived impacts of removing or reducing WAT use. Reflective, inductive thematic analysis [22] was adopted, allowing for deep insight into this complex and subjective topic to be gained.

### 2.2 Participants (selection and recruitment)

Participants were required to be aged 18+, English speaking, and previous full-time WAT users who had selectively reduced or completely stopped using their device. No restrictions on WAT types or WAT brands were placed in the current study, although examples of smartwatches, rings, and arm straps from brands such as Garmin, Apple, Fitbit, Coros, Oura, and Whoop were advertised in the recruitment poster. All levels of physical activity, types of WAT use, and participant demographics were welcomed in the current study to encourage wide perspectives in data.

The study was advertised by physical posters displayed around a northern university campus, alongside digital posters being advertised on social media platforms (specifically X, LinkedIn, and university news bulletins, and the local sports group pages).

Sample size was justified using Malterud et al.’s [23] information power. The study’s aims were clearly defined and specific in exploration, and participants were recruited on the basis that they were well-versed in the study’s topic. A strong, relevant theoretical knowledge underpinned the study, which informed the development of the interview guide and enabled the primary researcher to draw back upon to compare findings. Quality of dialogue was determined to be high within our transcripts, with rich, in-depth personal accounts shared by participants. Data was analysed rigorously through reflexive thematic analysis, extracting detailed accounts of participants lived experiences. Consequently, the research team deemed information power to be high, and were satisfied by a sample of fifteen [23].

The present study used Tracy’s [24] big tent criteria to strengthen the rigour of the data collection and demonstrates (a) a worthy topic (the research topic addresses a notable gap within the literature, by exploring both reasons for discontinuation but also behavioural and emotional responses from this decision), (b) research rigour (data was analysed in line with principles of reflexive thematic analysis, with justification for the decisions made), (c) sincerity (the research team acknowledged their experience and use of WAT’s), (d) credibility (data was triangulated within the research team as part of peer reflection), (e) resonance (participants quotes were rich in context which reflected the study’s aims), (f) significant contribution (the present study has encouraged a discourse about considerations in the ways that WAT’s are prescribed), (g) ethical (informed consent was obtained from all participants), and (h) meaningful coherence (the study’s aims, chosen methodology and analysis have clear cohesion).

### 2.3 Data collection

After electronically providing informed consent for the current study, interviews were arranged. Interviews were conducted by GH, SJ, and KM, either in-person (n=2) or online via Microsoft Teams (n=13). Interview durations ranged from 25.38 to 80.08 minutes (M=41.48, SD=13.72). Interviews were semi-structured, with prompts used to encourage conversation. Questions began gaining demographic information, before discussing reasons of device reduction or removal. Interviews then focussed on the perceived behavioural and emotional impact of reducing or removing a WAT. All interviews were recorded via Microsoft Teams and transcribed, with files automatically saved to a password protected personal file. These automated transcriptions were manually checked for accuracy and amended by GH and SJ.

### 2.4 Data analysis

Braun and Clarke’s six stages of thematic analysis were followed for data analysis [22,25]. GH, SJ, and KM independently familiarised themselves with data and began the coding process (stages 1 to 3) using Microsoft Word, before collaboratively reviewing codes and defining themes. A reflexive, inductive approach was adopted to encapsulate all data gathered. AG independently reviewed data transcripts and proposed codes to check data was accurately represented. All identified quotes were written into a table (see supplementary materials), with those most relevant discussed in the results section. Alongside this analysis, reasons for device discontinuation were compiled. These acted as an extension of a participant characteristics, rather than forming the thematic analysis, although participant quotes were added for further richness.

### 2.5 Reflexivity and research team

The positionality of the research team and how this may impact data interpretation were considered throughout the study, aiming for any personal biases to be minimised. The team consisted of four cross-disciplinary academics within psychology, sport and nursing domains, both male and female whose WAT use varied from no WAT experience (SJ), occasional WAT use (KM), and full-time WAT use (GH, AG, NS-H) to ensure diverse perspective. These factors were not conveyed to study participants to avoid social desirability bias in their reported answers. Interviews were conducted by GH and SJ whom are both academics in health subjects. Similarly, the positionality of GH and SJ was not revealed within interviews, aiming to minimise power dynamics and impact responses gained. While previous experiences with WATs will lead to some preexisting beliefs in the area, authors were cautious to only analyse the data presented on the page and not presume any other meaning. To further ensure parity, three authors first coded data and conceptualised themes (GH, GJ, KM), whereas two independently reviewed the transcripts (AG, NS-H) and table of quotes to ensure an accurate reflection of the data within proposed themes.

### 2.6 Ethical statement

Ethical approval was gained from the University of Salford ethics board in January 2026; reference 11716. Written informed consent was obtained from participants prior to arranging interviews, with agreement confirmed a second time prior to interviews beginning.

## 3. Results

### 3.1 Participant characteristics

The current sample consisted of fifteen participants (9 females, 6 males), ranging in age from 23 to 56 years (M = 35.33, SD = 10.57). All were involved in physical exercise at a recreational level. While participants were from a range of ethnic backgrounds, the majority of participants were White (White British = 5, White Irish = 2, White Scottish = 2, White American = 1). The majority of participants had fully removed their devices (n=10). Although, one participant within this category reported future plans to wear their device for a sporting event for communication purposes. Five brands were reported for discontinued devices (Apple, Garmin, Fitbit, Oura, and Coros), with Apple most popular (n = 5). Two participants reported last using smart rings, whereas the rest had previously used smartwatches. Before device discontinuation, WAT use had ranged from two to eleven years (M = 6.27 years, SD = 3.40. Duration of device discontinuation ranged from two months to ten years (M = 2.06 years, SD = 2.77). See Table 1 for full participant characteristic information.

**Table 1.**
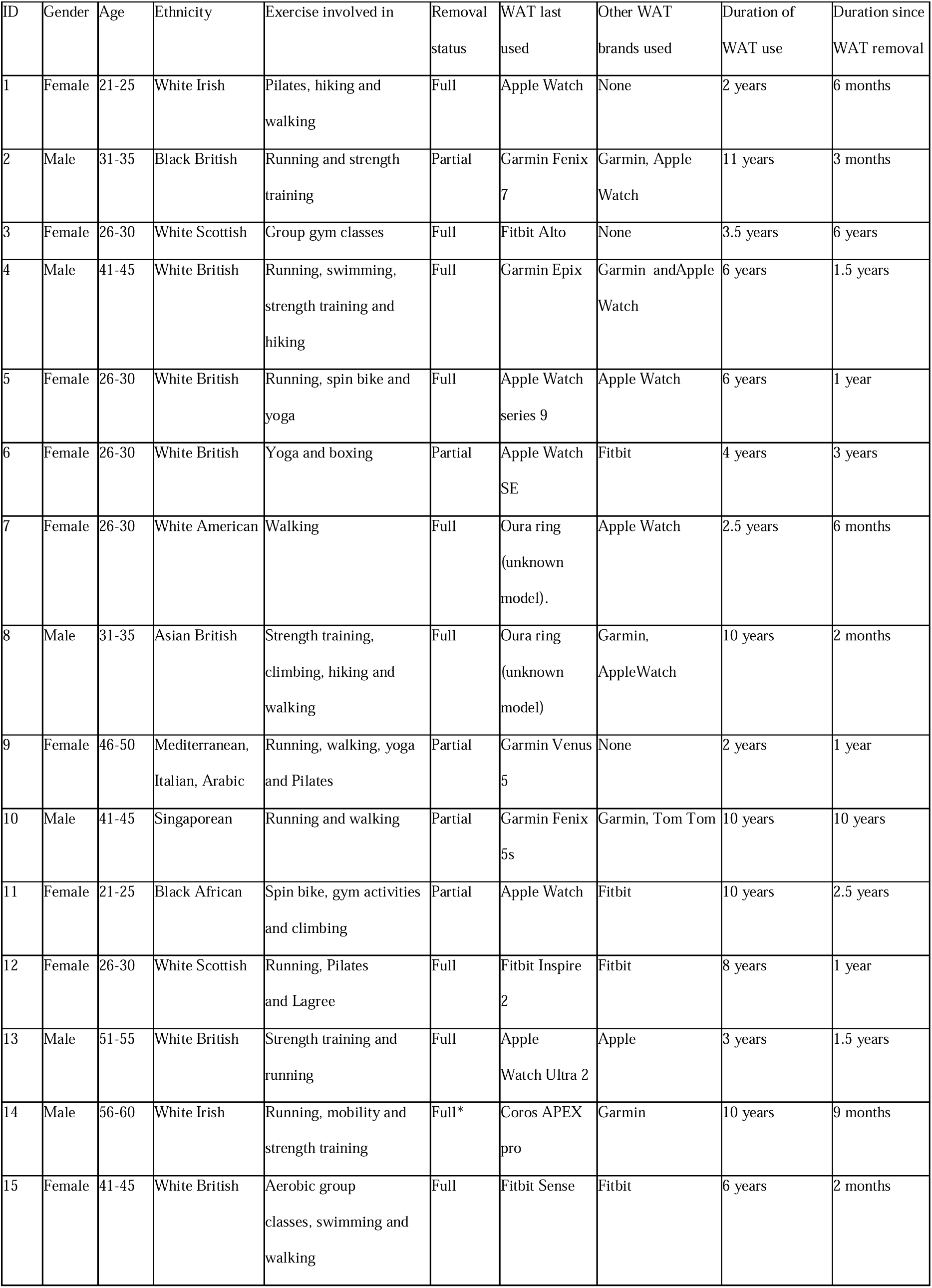
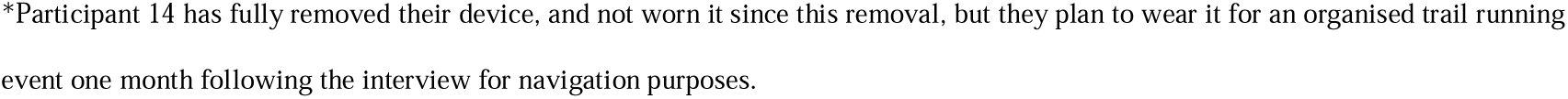
Participant characteristics.

| ID | Gender | Age | Ethnicity | Exercise involved in | Removal status | WAT last used | Other WAT brands used | Duration of WAT use | Duration since WAT removal |
| --- | --- | --- | --- | --- | --- | --- | --- | --- | --- |
| 1 | Female | 21-25 | White Irish | Pilates, hiking and walking | Full | Apple Watch | None | 2 years | 6 months |
| 2 | Male | 31-35 | Black British | Running and strength training | Partial | Garmin Fenix 7 | Garmin, Apple Watch | 11 years | 3 months |
| 3 | Female | 26-30 | White Scottish | Group gym classes | Full | Fitbit Alto | None | 3.5 years | 6 years |
| 4 | Male | 41-45 | White British | Running, swimming, strength training and hiking | Full | Garmin Epix | Garmin and Apple Watch | 6 years | 1.5 years |
| 5 | Female | 26-30 | White British | Running, spin bike and yoga | Full | Apple Watch series 9 | Apple Watch | 6 years | 1 year |
| 6 | Female | 26-30 | White British | Yoga and boxing | Partial | Apple Watch SE | Fitbit | 4 years | 3 years |
| 7 | Female | 26-30 | White American | Walking | Full | Oura ring (unknown model). | Apple Watch | 2.5 years | 6 months |
| 8 | Male | 31-35 | Asian British | Strength training, climbing, hiking and walking | Full | Oura ring (unknown model) | Garmin, Apple Watch | 10 years | 2 months |
| 9 | Female | 46-50 | Mediterranean, Italian, Arabic | Running, walking, yoga and Pilates | Partial | Garmin Venus 5 | None | 2 years | 1 year |
| 10 | Male | 41-45 | Singaporean | Running and walking | Partial | Garmin Fenix 5s | Garmin, Tom Tom | 10 years | 10 years |
| 11 | Female | 21-25 | Black African | Spin bike, gym activities and climbing | Partial | Apple Watch | Fitbit | 10 years | 2.5 years |
| 12 | Female | 26-30 | White Scottish | Running, Pilates and Lagree | Full | Fitbit Inspire 2 | Fitbit | 8 years | 1 year |
| 13 | Male | 51-55 | White British | Strength training and running | Full | Apple Watch Ultra 2 | Apple | 3 years | 1.5 years |
| 14 | Male | 56-60 | White Irish | Running, mobility and strength training | Full* | Coros APEX pro | Garmin | 10 years | 9 months |
| 15 | Female | 41-45 | White British | Aerobic group classes, swimming and walking | Full | Fitbit Sense | Fitbit | 6 years | 2 months |

\*Participant 14 has fully removed their device, and not worn it since this removal, but they plan to wear it for an organised trail running event one month following the interview for navigation purposes.

### 3.2 Drivers for device discontinuation

Participants reported eight key drivers for device discontinuation, including both full or partial removal of their WAT. See Table 2 for these drivers, alongside key quotes. An extensive table of all relevant quotes can be found in the supplementary materials (S1 Table).

**Table 2.**
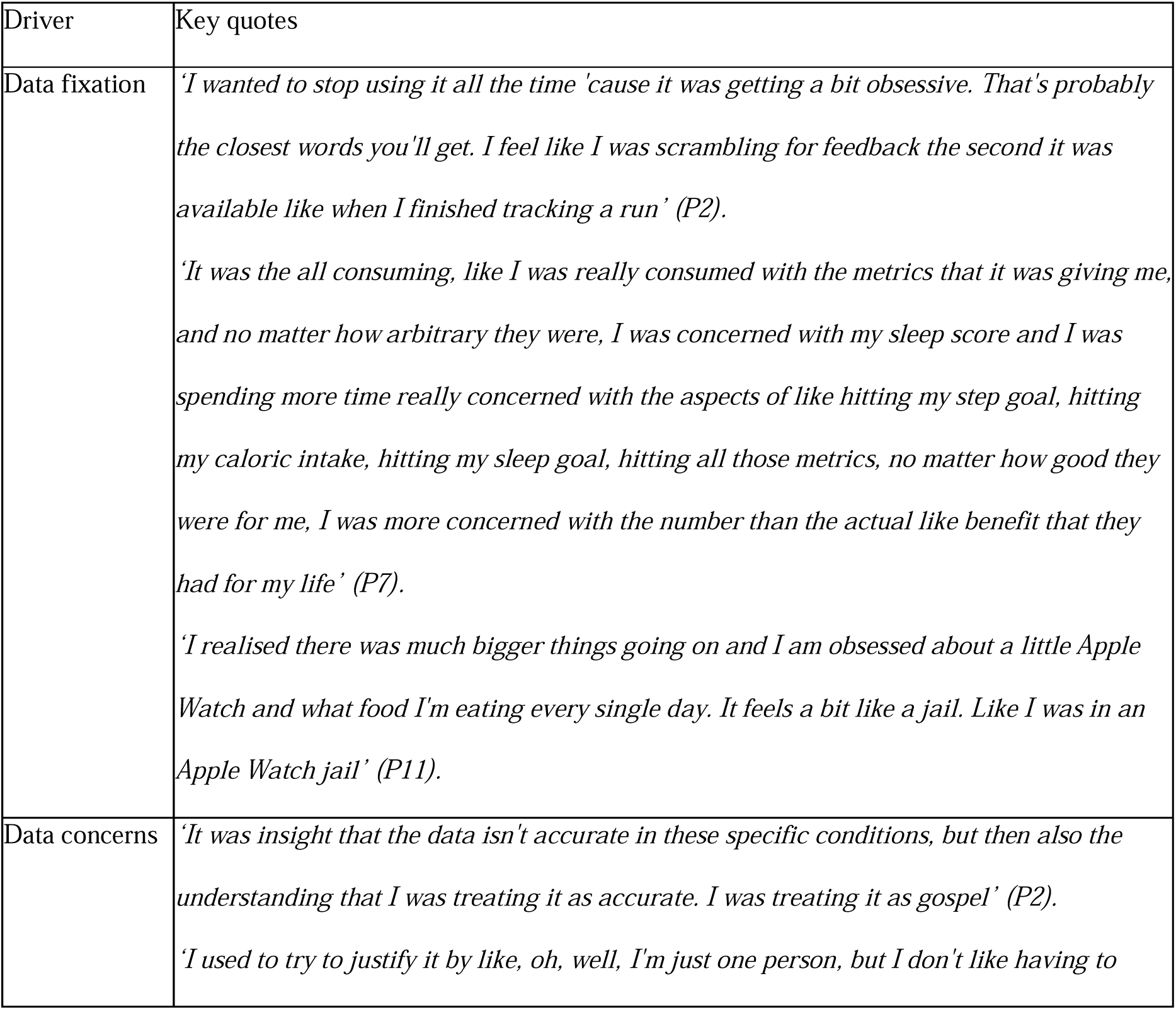
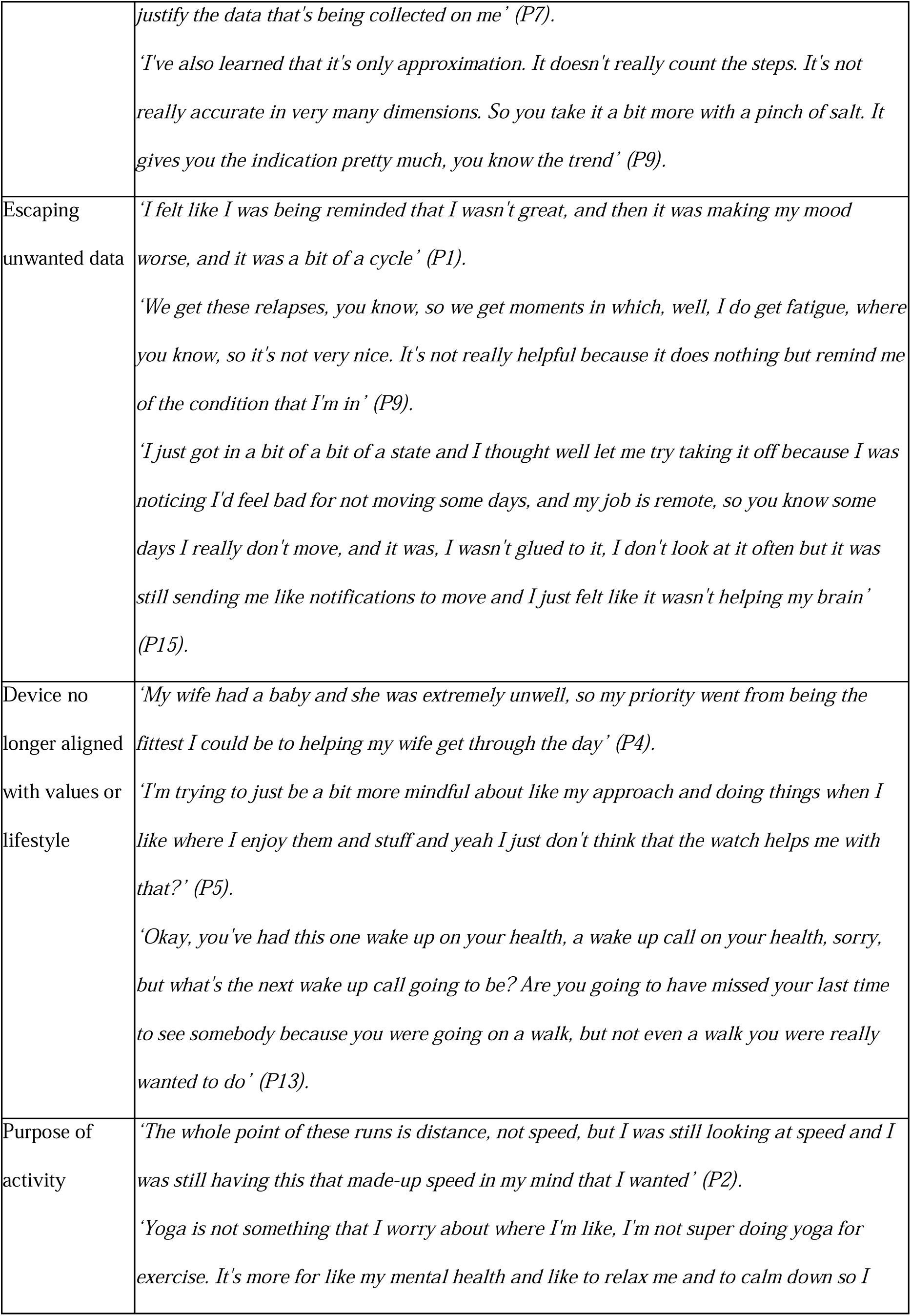
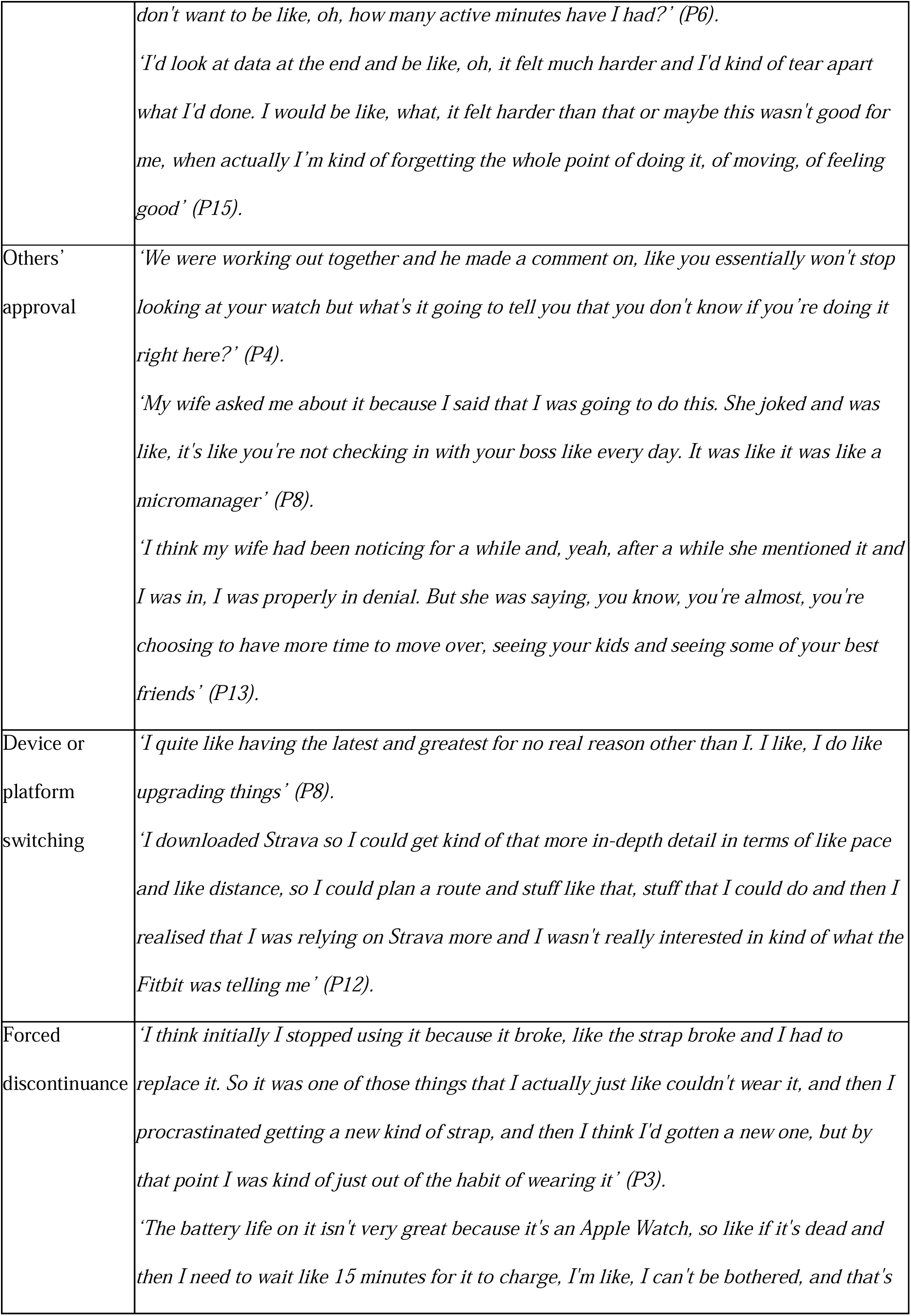
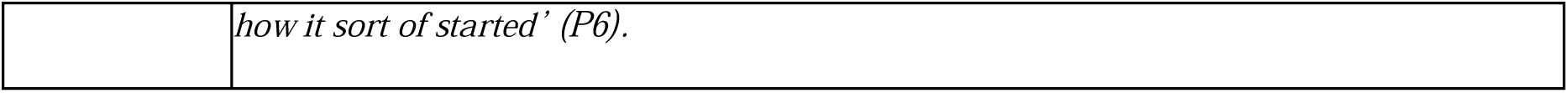
Drivers for device discontinuation.

The data a WAT provided was central to device discontinuation. Some users reported fixating on WAT data to an unhealthy degree, sometimes noticed by others, which led to device discontinuation. Whereas, other users reported concerns in both data accuracy or privacy, or the content of data, which led to device removal. Specifically, feedback perceived by the user as unhelpful led to discontinuation. This may have been emotionally unhelpful, for example, with users wanting to escape unwanted data, or practically unhelpful due to users having a lifestyle shift which meant exercise was no longer prioritised. A change in values around the body and the purpose of movement were also drivers for device discontinuation. Device switching was reported, with users switching tracking platforms or WAT models, resulting in removal of the previous device. Finally, some discontinuation was unplanned and forced due to technical issues.

### 3.3 Experiences of device discontinuation

Three themes were identified from collected data to illustrate the effects of WAT device discontinuation; Attitude change to activity, Reconnecting with the self, and Difficulties in habit reversal. Within these themes, nine sub-themes were identified. See Figure 1 for theme and sub-theme structure. The write up below discusses key quotes from participants; for a table of all relevant quotes see supplementary materials (S2 Table).

**Figure 1.**
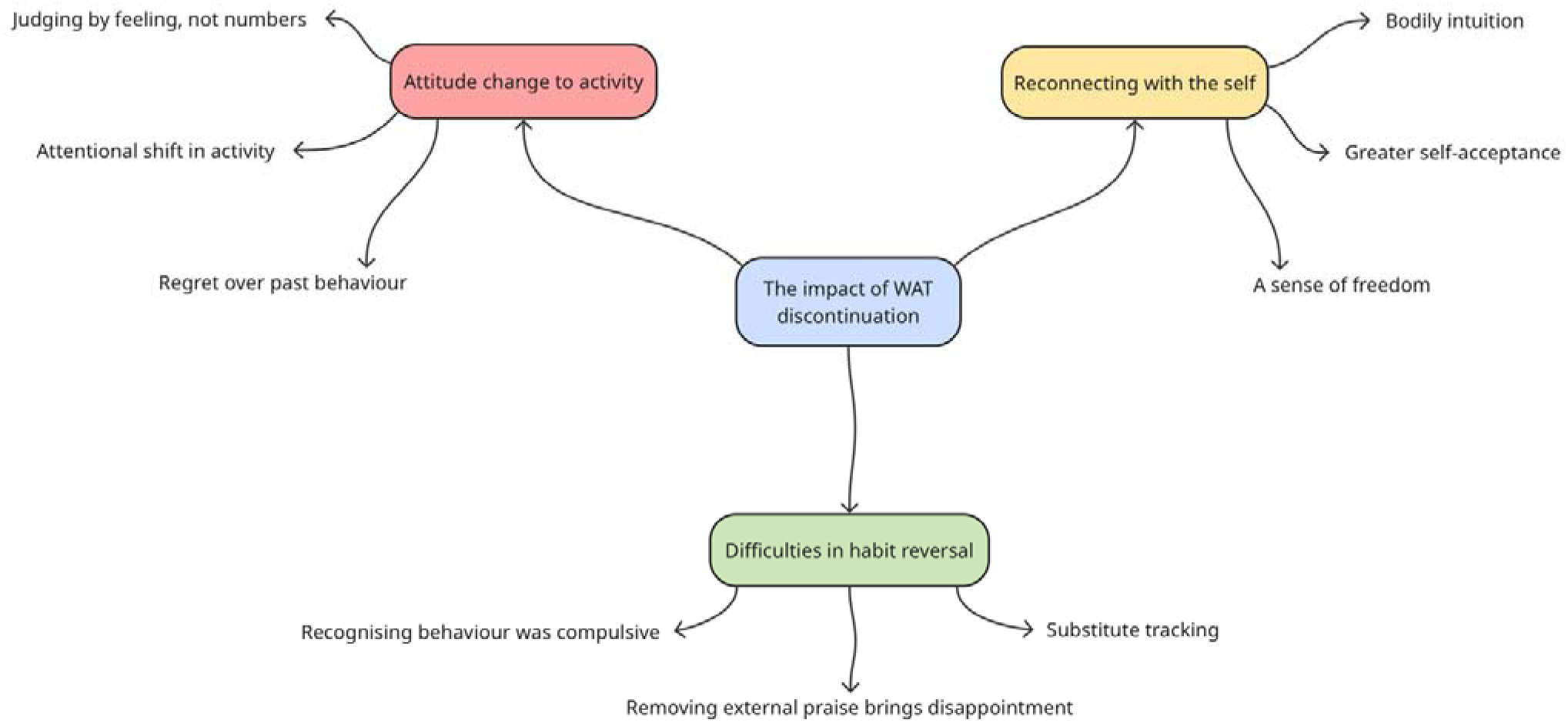
Thematic map.

#### 3.3.1 Theme: Attitude change to activity

This theme encapsulated a change in mindset participants experienced around physical exercise or general movement after device discontinuation. New beliefs around movement had formed including the purpose of movement, what ‘success’ is, and how this should feel. This theme was comprised of three sub-themes: Judging by feeling, not numbers, Attentional shift in activity, and Regret over past behaviour.

##### Sub-theme: Judging by feeling, not numbers

Participants reported device discontinuation due to previous fixation on WAT data, with one participant stating *‘I was dependent on it before, I just looked at it all the time’ (P4).* Consequently, they had grown reliant on the quantified and used data alone to determine their performance;

> *‘I thought how will I know if I’m getting better if I’m not wearing my watch and that was the panic, but also, that dependence on my watch to tell me if I was getting better and tell me how I’m doing essentially was the issue’ (P2)*.
>
> *‘I don’t compare my fitness anymore as a statistic, and I used to. It was purely about how fast I could do a 2K run or how many calories can I burn in a HIIT workout… I just thought about it in the numbers. It was never about my own actual health and fitness’ (P11)*.

This was also noticed by others, with a friend of the previous WAT user stating *‘you won’t stop looking at your watch but what’s it going to tell you that you don’t know if you’re doing it right here? (P4)* suggesting while the quantified self became the norm for WAT users, to those without a device, this seemed nonsensical and provoked confusion.

When using their WAT, participants viewed exercise based on its numerical impact;*‘I only burnt this many calories, so it can’t be useful’ (P1)* or *‘that was a good workout because I burned X amount of calories’ (P6)*. This meant participants were holding *‘an all or nothing’ (P7)* approach to exercise, viewing it only as a way to fulfil WAT goals which often resulted in negative emotion;

> *‘That made me feel a bit like shit, even during running these huge distances. That reminder this number’s gone down, this number’s gone up. It wasn’t nice. It was distracting me from the actual doing and the seeing, and making me forget you’re going to run, yeah, because it’s good for your fitness, but because you enjoy it’ (P14)*.

This suggests that by focussing on WAT data, participants ignored their own feelings about an activity, allowing numbers alone to determine whether exercise had been worthwhile. These set objectives left participants forgetting the emotional benefits of exercise, seeing movement only through the lens of data.

When device discontinuation occurred, participants no longer had access to the metric they used to wholly assess their exercise engagement?, meaning they were forced to change; *‘Now I’m having to judge what I’m doing from my body instead of my watch, which actually when you’re used to looking at pace and speed and distance is very different’ (P2).* Participant 2 added *‘I’m almost having to make peace with the fact that I can’t judge if I’m making progress on those long runs in that same objective way any more, but instead I can ask did I feel good?’ (P2).* This reflected that participants were having to consult their feelings during exercise now, but didn’t enjoy this reduction of numerical insight. Although, in time device discontinuation led to a changed mindset to exercise with pressure to meet numerical goals changing into a more relaxed attitude; *‘I think like the removal of it has really helped me to like reframe like, as long as I’m moving, that’s all that matters, I don’t need to do X amount’ (P7).* Now, participants must ask themselves *‘Do I feel at the end of the day like I did enough? Do I feel good in my body? Do I feel good in my mental health?’ (P7)*, and crucially, *‘Did I enjoy it?’ (P15)*.

Specifically, focussing only on WAT data seemed to reduce enjoyment during exercise, meaning device discontinuation brought about reflection upon previous WAT use. For example, Participant 13 stated WAT use ‘*Led me to forgetting the enjoyments of life, and movement, and connection. It became nothing to do with how I felt which is mad’*, suggesting a change in attitudes to exercise had come from device discontinuation.

Participant 11 had also reframed what successful exercise is, sharing;

> *‘I just wanted to burn as many calories as possible but now I know that that’s not the purpose of exercise. The purpose of exercise is 1) for health and 2) for your own mindset and wellbeing, so I’ll go because it feels good to move my body kind of vibe’ (P11)*.

Ultimately, participants reported that WAT discontinuation led to a shift in how they perceived success within exercise, moving from only using WAT data to judge categorically a success or failure, instead to seeing all movement as beneficial, and success deemed by feelings of strength and enjoyment.

Participants felt strongly about their previous judgement, sharing *‘I don’t think it’s very natural to have to have a machine tell you you’re doing a good job’* now believing *‘I think you should be able to find that within yourself’ (P4)*.

##### Sub-theme: Attentional shift in activity

No longer having WAT data to focus upon during exercise meant participants experienced an attentional shift. Participants found themselves focussing more on their physical surroundings during movement, which were previously not acknowledged due to attention placed on their WAT;

> *‘I was noticing daffodils. I was noticing signs of spring coming back and it was really nice and I know for a fact I wouldn’t have noticed that and I did enjoy it. You’re more connected with nature because you’re not connected to just another computer, really’ (P14)*.
>
> *‘You get used to staring at your wrist for a lot of it. I mean, it’s taken away from, like, just what you’re doing, the way you’re hopping along paths. The amount of views I must have missed or not fully taken in because I was looking at my wrist makes me pretty annoyed’ (P2)*.

This suggested participants felt more present within nature, taking in scenery which was previously not possible due to data fixation. In turn, this increased the enjoyment during movement; *‘I’m just like a mountain goat and I’m just trotting around, instead of before, I feel like I was trying to make things too efficient. I was being too rigid so I think it’s helping with enjoyment’ (P2)*.

Removing WAT data also brought a more rational viewpoint on health, seeing this as a bigger picture, over specific focus on daily elements. For example, Participant 13 said *‘I do think I’m just a bit more balanced. A watch could have told me one piece of information I wouldn’t know otherwise that would make me forget to acknowledge the rest of the workout, so just a bit more zooming out’*. Therefore, without WAT use, participants no longer had granular levels of feedback to focus on. Participant 7 echoed this change in focus, sharing *‘I was spending more time really concerned with all aspects of hitting my step goal, my caloric intake, hitting my sleep goal… I was more concerned with the number than the actual benefits they had’*, suggesting when using a device, focus was on meeting set goals by a device over both feeling and health benefits. Device discontinuation alleviated feelings of panic from Participant 5 who said *‘It’s helped me be a little more relaxed about things I think, and possibly have a bit more of a long term view [on health]*’.

##### Sub-theme: Regret over past behaviour

Furthermore, device discontinuation encouraged participants to reflect on their previous use. Participant 14 stated *‘I had just become a bit of a robot I think’*, boldly highlighting their unnatural focus on data and mindset on movement.

Regrets were typically around how much importance users had placed in their devices. Some shared mild regret about their use, for example, *‘With 10 years of hindsight I can tell you that it probably wasn’t good. I think, I just used it for everything and I got so into it’ (P8)* showing how much users listened to their WAT data. These set goals also led to participants forming negative attitudes around resting. For example, Participant 1 shared *‘I found myself really beating myself up when I got busier with a job and and when when I got ill’*, suggesting these goals set by a WAT didn’t accommodate changes in availability and ability to exercise.

While some regret was mild, others reported high levels of regret;

> *‘You’ve had one wake up call on your health, but what’s the next one going to be? Are you going to have missed your last time seeing somebody because you were going on a walk, but not even a walk you wanted to do, it was wanting to get your steps in’ (P14)*.
>
> *‘She was like, I kind of need you to be there because something’s happened… Her best friend had died and she got that information. And I wasn’t there because I wanted to go on a walk to get my steps in, which was so unnecessary. I didn’t need to do that’ (P11)*.

Heightened emotions were reported in these participants, sharing ‘I have a bit of hatred towards it sometimes when I look back’ because ‘getting a device age 13, 14 into adulthood really did take a piece of my childhood away’ (P11).

Specifically, Participant 11’s regret came from the unhealthy behaviours their WAT device facilitated; *‘I really don’t think I would have had an eating disorder if it wasn’t because of it’* showing the severity of behavioural impact that people can face from WAT use.

Participant 13 detailed similar extreme regret after their wife noted the consequences of their WAT use;

> *‘She was saying, you’re choosing to have more time to move over seeing your kids and seeing some of your best friends and, yeah, I think I was moaning at one point about like going on holiday with two of my best friends because they like to relax… and that broke me a bit because, well, they’re my best friends! I love them to piece and I was genuinely hand on heart not impressed to be spending time with them because I’d get less steps in that day. I fills me with anger now or, or even a bit of shame’ (P13)*.

Data highlighted varied levels of regret in participants, with some reporting none, whilst others reporting high levels. Across quotes, data illustrated that WAT use had led to time spent focussing on their device and meeting set goals, which left social relationships compromised.

#### 3.3.2 Theme: Reconnecting with the self

Device discontinuation also brought about a change in mindset around the self. The removal of WAT data resulted in participants adopting a less critical view of themselves and reconnecting with their bodily intuition around when and how to exercise. This theme was comprised of three sub-themes; Bodily intuition, Greater self-acceptance, and A sense of freedom, illustrating psychological and emotional impacts of device discontinuation.

##### Sub-theme: Bodily intuition

Participants reported using a WAT to determine when to exercise, with data being placed over their own bodily intuition around energy levels or motivations. Participants 1 shared *‘I was looking at a screen instead of asking myself how I felt’* suggesting data were placed above one’s feelings. Having data led to an inflexible view on exercise routine; *‘before I was almost thinking of it as like this, this employee that had this to-do list every day and like had to do it’ (P8)*. Participant 4 echoed this, saying they felt *‘like test subject or like product’*, suggesting WAT use led to self-objectification and activity feeling non-negotiable in users. Due to this, device discontinuation was difficult to navigate for participants;

> *‘I had to learn to trust my body again, which sounds sounds huge, doesn’t it? But I got so used to looking at a screen when I took it off I was I was questioning like how am I going to do this and how will I do this?’ (P1)*.

This highlighted participants had to relearn their wants around physical exercise after their device had led them to ignore these. This left some users stopping physical exercise completely after reevaluating their motivations;

> *‘I stopped like exercising… I just don’t think I like wanted to, or in the headspace I was in, and the kind of part of life I was in, I didn’t feel pressured to do it anymore. So I was like, I’m just going to do what I want’ (P3)*.

Whereas, before device discontinuation, decisions around exercise were overly influenced by WATs; ‘Before it was like I was making decisions for a collective or me and the watch, which sounds stupid, but numbers were always in mind’ (P14). This suggested a shared goal had formed between user and WAT with blurred boundaries between the two. Participant 9 echoed this, reported that ‘Since [device discontinuation], now I’m in control. I’m not being a slave to the watch’, again, highlighting the control a WAT had over participants’ behaviour.

This gained bodily intuition around movement has resulted in participants feeling more relaxed;

> *‘I feel like I’m probably just a bit more relaxed. It’s not that I’m being lazy, but I’m weighing up like, should I do this workout today or tomorrow? And I’ll be like, oh, I’m quite tired today. Whereas I would have looked at data before and said I could do with doing that now’ (P8)*.

This suggested that bodily intuition was being ignored in order to achieve thresholds set by WATs, which users felt pressure to fulfil. Therefore, device discontinuation left participants listening to their body before deciding if to exercise; *‘I’ll go and exercise when I want to go and exercise. And I’m doing it for me and my own health and fitness, not because a watch is telling me that I’ve only burnt so many calories today’ (P11)*. For many, the exercise itself didn’t change, but frequency of movement and motivations did; *‘It’s more like the mental processes behind it and you know, why am I doing it? Do I want to do it or is it just on my schedule today? If I don’t want to do it, can I swap it round?’ (P13)*.

For some, realising they had lost bodily intuition around exercising was a shock. Participant 1 said *‘I thought I was looking into my body more but I was actually looking at a screen’,* suggesting users typically expect to learn more about their body from getting a WAT, yet instead, end up ignoring their body. Therefore, device discontinuation resulted in participants feeling more connected to their own bodily intuition.

##### Sub-theme: Greater self-acceptance

Device discontinuation led to a greater self-acceptance, both in terms of movement and body image, in participants. This occurred because exposure to WAT data was having a negative emotional impact on participants; *‘having that data, I just started being so critical’ (P1)*. This impact was also noted by participants’ peers, with Participant 4 sharing *‘he said, seriously, I don’t think I’ve seen you look at that watch and look happy from whatever it’s telling you’*. This showed that WAT use was resulting in negative mood, which in turn was impacting others around them.

WAT data allowed users to fixate on metric and assign worth to these values;

> *‘Some days might be quieter and some days might be more active, and that’s the ebb and flow of the working week… Tired isn’t bad, but I saw it as something to deny and ignore when I was wearing my watch’ (P13)*.

Here, device discontinuation left Participant 13 feeling more positive about days with low levels of moment, highlighting how resting was viewed negatively during WAT use. This was also reported by Participant 3; *‘If I didn’t have an active day I’d feel rubbish’*, again, suggesting that WATs encouraged every day to be active. A kinder attitude was reported around exercise also. Participant 15 stated *‘I feel a bit less, like, frustrated by myself, I would beat myself up some days for essentially being bang average, when average is normal’*. Here, this numerical judgement of data promoted a critical view of the self, with users feeling disappointed by their average performance. While WAT data was not being overtly negative, WATs facilitated this critique; *‘It was me doing the bullying, saying this isn’t good enough, but it was my watch whispering the things in my ear to say out loud’ (P14)*. This highlighted that WATs provided data which may be harmful in vulnerable populations. Alternatively, device discontinuation meant participants no longer had metrics to use for negative self-talk.

Other participants reported that device discontinuation had led to greater self-acceptance around their body image;

> *‘If I think about my like watch usage, it’s always been connected to my own like body image and things like that and like activity as a way of like managing that, I suppose. And I think, yeah, so I think that like process of taking it off is kind of, for me, part of like a more intentional control for just like trying to change how I think about these things’ (P5)*.
>
> *‘I do struggle a little bit like with like my weight and things like that, and so I think it was just a way for me to like kind of bully myself further… it was a way to like put a numeric number on it, like, oh, you are this way because you didn’t hit this metric. And so I think removing it has sort of like, I’m very neutral about those things now’ (P7)*.

This suggested that WAT data led to continuous negative thoughts around body image, whereas device discontinuation led to body neutrality. Participant 5 supported this idea, stating *‘I just feel like I have a more pragmatic view now’ (P5).* Although, they struggled to identify whether their device discontinuation led to greater self-acceptance around movement, or whether this self-acceptance prompted them to remove their device, stating;

> *‘I think me trying to like be a bit easier on myself kind of led to not wearing the watch as much. So I can’t tell how much not wearing the watch has like enforced that. I do think it probably has helped, yeah, there were just some times where I would just feel so, just so irrationally like down and hard on myself because the number on this like stupid watch wasn’t right’ (P5)*.

This linked to some participants discontinuing device use due to the WAT no longer being aligned with their values, suggesting that greater self-acceptance may be both an outcome of and driver for device discontinuation.

##### Sub-theme: A sense of freedom

Prior to device discontinuation, a human-computer interaction had left participants no longer thinking independently about their behaviour;

> *‘It does become this combined mindset, really. It’s not you versus you watch, it’s like you’re a team, you’re on the same team and wanting to get better data. Removing that has meant it’s just me, so I’m seeing myself a bit more as a just normal person instead of a machine’ (P14)*.

This illustrated the fact that participants were now making decisions with the aims to please their WAT over their own motives, recognising now the issues with this. Given the glanceability of many WATs and typical full-time wear, *‘It was a constant back and forth on numbers on my brain’* added Participant 14. Participants similarly shared *‘It slowly consumed my life’ (P7)* and that feedback was *‘heckling’ (P4),* highlighting the repetitive nature of feedback and the extent it impacted users. Therefore, removing this constant occupation around data led to participants feeling a sense of freedom;

> *‘It definitely felt more kind of freeing…I think it did change the way I view exercise a little bit and I don’t think that it effected how much I exercise. I think I was able to be more consistent, but kind of less dreading doing stuff’ (P12)*.

This suggested that participants felt under surveillance when wearing a WAT. This feeling of restriction being lifted via device discontinuation was echoed by Participant 11 who compared their WAT use to being incarcerated;

> *‘I realised there was much bigger things going on and I am obsessed about a little Apple Watch and what food I’m eating every single day. So that’s when I kind of, I don’t know how to describe it. It feels a bit like a jail, like I was in an Apple Watch jail’ (P11)*.

Therefore, when a participant had discontinued WAT use they reported feeling ‘so relaxed’ (P8) and ‘completely peaceful… because I don’t know if you can fully switch off when you’re looking at your watch that regularly’ (P2) again highlighting that WAT use resulted in stress. Specifically, Participant 4 said ‘It felt like I’d put my phone on silent mode and I’d like I blocked it out, I put like noise cancelling headphones in’ (P4) suggesting device discontinuation brought a feeling of escape. Now, this had allowed participants to slow down and adopt a different lifestyle;

> *‘They just made me a bit too obsessed with getting better all the time … I’ve started trying to have like a slower life at a slower pace and and like more relaxing and I think taking it off is something that’s really helped me’ (P1)*.

This gained sense of freedom left participants enjoying exercise more. As mentioned, Participant 4 likened themselves to *‘A mountain goat and I’m just trotting around’ (P2)*, implying they feel present in their surroundings.

Similarly, Participants reported *‘I don’t worry as much about like, oh, how, like, what’s my heart rate going to be just now? Or am I expending enough energy like doing this exercise? (P6)* and this *‘removed a lot of the kind of anxiety I felt about like not getting better all the time’ (P8)*.

This sub-theme illustrated the constant provision of feedback participants felt during WAT use, meaning device discontinuation led to a quieter and more relaxed life. This was both applied to exercise and daily activity, which resulted in a feeling of freedom now their WAT isn’t *‘shouting in my ear the whole time’ (P14)*.

#### 3.3.3 Theme: Difficulties in habit reversal

While device discontinuation brought positive emotional and psychological impact, participants also faced difficulty in habit reversal. Specifically, WAT use had become a compulsive behaviour for participants meaning device discontinuation was challenging. Participants had determined exercise value via WAT data, meaning new ways to judge exercise had to be learned. Additionally, this removal of data left some participants finding alternative methods to monitor behaviour. The theme consisted of three sub-themes; Removing external praise brings disappointment, Recognising behaviour was compulsive, and Substitute tracking.

##### Sub-theme: Removing external praise brings disappointment

Participants reported that WAT feedback often made them feel positive about their workouts;

> *‘I loved it. It was this cheerleader and it’d be like yeah, you’re doing amazing! Like, keep going! It was really good for me and I think now I’ve stopped using it I don’t want to admit how much I did really enjoy that, but I did’ (P1)*.

Therefore, device discontinuation left participants missing this praise; ‘It would it would be nice to know what I’d done’ (P4), ‘I’ll see people tracking things and every so often without sounding stupid, I do feel a little bit envious’ (P13). This highlighted that device discontinuation led to participants feeling like they were missing out, both on praise and insight into their behaviour. Therefore, they needed to navigate these negative emotions to maintain discontinuation.

Participants seemed to specifically miss WAT feedback when they perceived their performance as strong; they were *‘so disappointed because I didn’t have the watch telling me I’d done good’ (P1)*. This feeling arose from believing WAT data confirmed their performance and provided a level of validation that doing alone did not provide;

> *‘It feels like you’ve got no evidence of what you’ve done. It’s really hard to explain it to someone that hasn’t used a device… your recordings become proof that you’ve done something, it becomes like a little logbook. And so, when you have all of those, and then you stop, it does feel like you’re not gaining the same thing from it… it felt really different and I think it’s what I hated the most about removing it’ (P13)*.

This demonstrated that exercise with and without a WAT felt psychologically different for participants, with WATs acting as proof of performance to users. This links to the quantified self [10,11], with participants still seeking for numbers to represent their experiences over their own feelings and perceptions.

Device discontinuation also restricted participants from receiving peer feedback. Strava [26] is a social media platform where users can share their exercise and receive feedback. Without WAT data, users could no longer share detailed activity with their peers;

> *‘Strava was funny because you’re showing people what you’ve done. I don’t want to say it’s boasting, but it is drawing attention to you. It’s saying, look at what I did this Sunday morning, and so it was a bit of a second layer of reward, like people giving you kudos, they’re being like, yeah, fair play like this looks ace smashed it. You are removing some reward’ (P2)*.

Therefore, device discontinuation left participants missing out on feedback from both their device and peers, now navigating exercise without praise.

##### Sub-theme: Recognising behaviour was compulsive

Many participants reported their data fixation led to device discontinuation. Participants were checking their WAT excessively; *‘You’re just constantly checking your heart rate, you’re checking how many calories you’ve burnt, you’re checking how much activity you’ve done every like 2 minutes’ (P11)* due to the accessibility of it; *‘If it’s there, I’ll look at it’ (P14).* Participants tried to change this behaviour previously by reducing data access whilst still wearing their WAT, but reported difficulty;

> *‘The data was there so I couldn’t help but look at it, which sounds like no self-control, but actually when it’s all in front of you to like such detail, it’s hard not to look at it. So I looked at everything, I really did’ (P1)*.
>
> *‘Before I took it off, I felt a bit like, silly, I thought, oh, you can just ignore it, but if it’s there on an app, I’m not ignoring it… it felt like a weird self-control issue with me, and I did feel a bit silly’ (P8)*.

This highlighted the compulsivity of this behaviour, with participants struggling to resist urges to look at their WAT despite their efforts. Furthermore, Participant 11 likened their WAT use to an addictive behaviour;

> *‘I was just trapped in it and the only way I can compare it is, you know when you hear about how if people have a problem with alcohol and they go to maybe different petrol stations every day, so people don’t know that they’re drinking as much and they hide things under their bed, that kind of thing? I feel like that was me with my Apple Watch’ (P11)*.

This demonstrated the severity of problematic WAT use faced by some participants, and the secretive nature adopted during this behaviour, with participants potentially feeling embarrassed or confused over why they could not control their WAT use.

Participants reported that compulsive WAT use specifically occurred when they had completed daily tasks (e.g. step count and active minutes reached) multiple times, with a streak forming. When participants had done so for longer, a greater consequence was felt if they lost this progress; *‘the higher the number, the more you don’t want to get rid of it’ (P8)*. This meant behaviour felt non-negotiable for participants with pressure felt to continue this streak;

> *‘If you didn’t do this 3000 steps today, you could physically do it tomorrow, you could catch up but it was having this little sticker on a calendar chart that just meant to me that if I didn’t do one, it was this glaring, it was almost a big circling, so you know, you missed out here. And they work on streaks on Apple, so the longer you did it, the more obliged you were to carrying on’ (P13)*.

Given WAT use is typically continuous, participants did not realise the compulsivity in their behaviour until they resorted to device discontinuation. Participant 11 stated *‘I didn’t tell them that I was obsessed with it because at the time I didn’t even realise I was obsessed with it’ (P11),* highlighting this WAT use occurs gradually, leaving users unaware of their problematic behaviour.

Specifically, this type of WAT use was common; *‘using my watch like this was just like the normal thing to do’ (P4)*, and when participants told others their plans of device discontinuation they were greeted with shock; *‘they were like, oh my God, I would die without my Garmin or my Apple Watch’ (P12)*, implying that device dependency and compulsive data use is seen as typical WAT use in society.

The compulsivity of this behaviour meant habit reversal was extremely difficult. Participants experienced heightened emotions, reporting *‘I was itchy, I was irritated, I hadn’t really felt the benefits yet, I was just missing my watch’ (P13)*. These are symptoms in line with those trying to reverse behaviours recognised as addictive, demonstrating the negative emotions faced during behaviour change. Participants were required to navigate these emotions and relearn how to go about their lives without a WAT, which was a stressful experience;

> *‘I had this full, like, adjustment period in crisis because I was so used to looking at my watch… I was so connected with it all the time. I was checking my wrist so often and every time going, oh God, it’s not there, how am I meant to know this? So it was weird. It was genuinely a 180 when I took it off. I must have been looking at it so often cos I just remember feeling lost for a while’ (P13)*.

This quote demonstrated long-term routines that had been established around WAT use, with participants not knowing what to do without it, and the change in mindset that participants had to navigate.

##### Sub-theme: Substitute tracking

Device discontinuation meant participants no longer had recording of their activity. This was missed by participants, so alternative methods of logging their exercise were found.

Some participants opted to manually track their exercise on pen and paper. For example, Participant 13 shared *‘I’ve got a tiny notebook that I shove in my gym bag and I just make a note of where I left off at to see do I need to go up away or should I stick on this?’ (P13)*, implying that keeping a record of activity helped them progress. Similarly, Participant 2 has *‘A little list of what I do on my long runs that I’m physically writing down in a notepad, so I do know what I’ve done’ (P2)*. This also suggested that recorded data allowed for participants to have a record of their past activities, acting as a memory prompt over a longer period of time.

Strava was used by Participant 12 for this same reason;

> *‘I still put in like a yoga class onto Strava and it’ll just be one hour yoga or one hour Pilates, but it’s got nothing to do with like the amount of calories I burned or anything like that. Like it’s just not that important to me anymore. And so now it’s literally just to keep track of like, say, an average of four things a week’ (P12)*.

While WAT data was no longer collected, limiting the metrics which would be posted online, Participant 12 still wished to post on this platform for their own personal recording of workout frequency, suggesting that this recording helped them maintain a routine.

Whereas, others resorted to asking their friends for WAT feedback; *‘A couple of times I’ve been with my friends and I have asked like essentially, what we’ve done from from their watch, just to, just to get a bit of that data back’ (P4)* highlighting that participants still wanted access to this data despite the reasons for their device discontinuation. This was also reported by Participant 8 who said *‘I can ask someone else what we did if we’ve been walking together, like if I really want to know’,* illustrating again that some participants still wished to receive data. Therefore, asking to see others’ data allowed this, without participants reversing their own device discontinuation and negating the positive impacts experienced.

## 4. Discussion

### 4.1 Summary of findings

Three themes with a total of nine sub-themes were identified from the dataset. Specifically, no longer having WAT data left participants having to judge their performance based only on feelings during exercise. While participants now felt more present during their workout, this difference brought feelings of regret over past behaviour. This lack of WAT feedback also reduced feeling pressured to exercise, with a new sense of freedom expressed by participants. Additionally, participants were now less critical and more accepting over their bodies and fitness.

Although, device discontinuation also negatively impacted participants. Participants missed the external praise and confirmation of activity that their WATs provide, with some finding alternative methods to track this.

Furthermore, navigating device discontinuation was difficult for users, with many experiencing heightened emotions such as stress and irritability following this action. However, these feelings were reported to subside after an adjustment period.

### 4.2 Existing literature

Existing literature identified changes in health status, activity level, life circumstances, feeling discomfort around data, and no longer wishing to receive data as drivers for device discontinuation [16–18]. The current study supported these findings, with these factors leading to device discontinuation. Furthering this, the current study also identified explanations that had not previously been identified within literature to our knowledge, including device discontinuation due to noticing their fixation on WAT data, others’ lack of approval on their WAT use, or technical issues. Elements of gamification are used by WATs to promote user engagement [27]. Specifically, participants detailed being impacted by ‘streaks’ being built up over longer periods of time which led to data fixation. While this gamified element had led to continued interaction with WATs, supporting findings by Lima [27], exercise no longer felt optional and this negatively impacted participants. Therefore, the current study highlights the dangers in encouraging repeated WAT use, with possible outcomes of exercise addiction, eating disorders, or impaired mental health and relationships.

In line with other literature, the quantified self had occurred in users prior to device discontinuance, resulting in a loss of bodily intuition around exercise [6,10,11]. These findings extended our current knowledge of the quantified self by showing this mindset was reversible over time via WAT removal. This reversal of the quantified self, facilitated via WAT discontinuance, specifically improved exercise enjoyment and reduced distraction, in line with findings by Tapia [19]. Furthermore, Tapia [19] reported conflicting findings with some participants reporting they missed receiving data. Current findings echoed this with participants simultaneously appreciating no data provision, yet missing data provision. The current findings provided further insight into why some users missed receiving data, identifying that this removed both a level of reward from exercise and a level of confirmation that an activity had been completed.

The idea that exercise has more value if recorded suggested that some participants remained impacted by the quantified self mindset [11,12], even following device removal, highlighting the long-term impacts of WAT use. These findings also suggested that WATs promoted increased physical exercise via increasing extrinsic motivation, supporting existing literature [7,8].

Although, given participants reported feeling freedom and relaxation following device discontinuance, this questions whether exercise was completed because of motivation or pressure; a debate which also occurred in our previous research [6].

Additionally, the current study identified new learnings around WAT use and device discontinuance. Specifically, participants detailed regret over their previous WAT use which only occurred after device removal allowed for a period of reflection. Regret was reported specifically around neglecting relationships to prioritise achieving positive WAT data, and how individuals treated themselves. While no study has directly examined the impact of WATs on social relationships, this may relate to phubbing - the concept of snubbing individuals in social settings to instead look at one’s phone [28]. While this terminology specifically applies to being invested in one’s smartphone, many WATs now have smartphone features, meaning this may be applicable within this setting. Furthermore, phubbing has been linked to reduced mental health, impaired relationships, and device dependency [29] which were all reported as drivers for device discontinuance in the present study.

The current study found that device discontinuance led to greater self-acceptance within participants, both in terms of body image and exercise. Findings supported previous literature which identified tracking apps as significantly linked to negative body image and restrictive eating [30], with individuals whom track their behaviour often adopting the mindset that food should be earned via exercise [31]. Similarly, Participant 11 in the current study explicitly believed that using a WAT is what led them to develop an eating disorder which spanned over a decade. While findings within this study and over large spanning reviews, shows that WATs lead to an increase in physical activity [5], this may be at the detriment of one’s mental health. Viewed through the public health principle that there is no health without mental health [32], this evidence questions the long-term impact of WATs on individuals’ mental health, and, in turn, whether WATs can be labelled as health promotion tools.

### 4.3 Strengths, limitations, and implications

As with all research, the current study has limitations. Participants shared their experiences of device discontinuation over a vast duration and had used a range of brands, the majority of participants were white females. While this population is consistent with the typical demographic of WAT users [33] and reflects wider challenges faced in recruiting unrepresented groups in research [34], the limited diversity within this sample may hinder the transferability of our findings. Additionally, participants reported their subjective reports on changes in well-being and exercise performance over a long period of time.

Accounts may have been difficult to remember and/or narrate to researchers, particularly as many were detailing large shifts in their beliefs, meaning data may be somewhat limited or not truly representative of long-term experiences. Although, information quality of data collected was high. Finally, length of WAT use and duration following device discontinuation was extremely varied between participants. While one could argue this provided a range of insights into these experiences at different time points, this lack of standardisation within use and discontinuation may limit comparisons between participant experiences. The current study provided new learnings on WAT discontinuation and filled an existing gap in literature, although the conduction of diary studies tracking participants both pre- and post-WAT removal could be insightful future research within this area.

These results hold vast implications around what safe WAT use looks like. While device discontinuation occurred for a variety of reasons, many participants had become fixated on their WAT data which had impacted their mental health. Online safety is an increasing policy focus in the UK reflected in the 2023 Online Safety Act [35]. Yet, despite concerns regarding digital wellbeing, there is currently no formal guidance defining healthy WAT use or identifying whom may be at risk of harm from WAT use. Developing such recommendations is crucial as the NHS begin to incorporate these devices as healthcare promotion tools [36,37].

Furthermore, the current study held implications around behaviour change promotion. WATs have been shown promising tools for exercise promotion [5], although, the current study found that this same WAT use may be impacting individual’s mental health and promoting unhealthy attitudes to exercise. While we must promote exercise uptake to improve health outcomes, this should not be at the detriment of mental health. Therefore, future interventions in this area should aim to promote exercise, while promoting positive well-being and enjoyment in movement in order to bring long-term physical, mental, and social health improvements.

## 5. Conclusion

The current study provides novel insight into the experiences of WAT discontinuation amongst former users. While WATs are commonly use to increase physical activity, findings demonstrated that device discontinuation often resulted in greater self-acceptance, enjoyment of movement, and reduced pressure surrounding exercise. This highlighted that WAT used shapes how users’ perceive their bodies, exercise and self-worth, therefore, device discontinuation led to change attitudes. However, participants also faced feelings of frustration, irritability and loss following discontinuation, which they had to navigate before feeling the positive effects of device removal. While these feelings subsided over time, some users reported difficulty having to navigate these.

Overall, the findings highlight the complex relationship between behaviour change and well-being, which WAT use and discontinuation impacts. Although WATs may successfully promote physical activity, their long-term impact on users’ mental health requires greater consideration.

## Data Availability

All data produced in the present study are available upon reasonable request to the authors.

## Notes

### Competing Interest Statement

The authors have declared no competing interest.

### Funding Statement

This study did not receive any funding

### Author Declarations

Ethics committee of University of Salford gave ethical approval for this work.

## References

1. An HS, Jones GC, Kang SK, Welk GJ, Lee JM. How valid are wearable physical activity trackers for measuring steps?. European journal of sport science. 2017 Mar 16;17(3):360–8.

2. Kumar N. Smartwatch statistics (2025) – Users & market share. DemandSage. 2025. Available from: https://www.demandsage.com/smartwatch-statistics/

3. Henriksen A, Haugen Mikalsen M, Woldaregay AZ, Muzny M, Hartvigsen G, Hopstock LA, Grimsgaard S. Using fitness trackers and smartwatches to measure physical activity in research: analysis of consumer wrist-worn wearables. Journal of medical Internet research. 2018 Mar 22;20(3):e110.

4. Shin G, Jarrahi MH, Fei Y, Karami A, Gafinowitz N, Byun A, Lu X. Wearable activity trackers, accuracy, adoption, acceptance and health impact: A systematic literature review. Journal of biomedical informatics. 2019 May 1;93:103153.

5. Ferguson T, Olds T, Curtis R, Blake H, Crozier AJ, Dankiw K, Dumuid D, Kasai D, O’Connor E, Virgara R, Maher C. Effectiveness of wearable activity trackers to increase physical activity and improve health: a systematic review of systematic reviews and meta-analyses. The Lancet Digital Health. 2022 Aug 1;4(8):e615–26.

6. Humphreys G, Jensen S, Gluchowski A. “It’s like a toxic relationship”: Examining internal conflict experienced in wearable activity tracker users. PLOS Digital Health. 2026 Feb 20;5(2):e0001136.

7. Jung EH, Kang H. Self-determination in wearable fitness technology: the moderating effect of age. International Journal of Human–Computer Interaction. 2022 Sep 14;38(15):1399–409.

8. James TL, Wallace L, Deane JK. Using Organismic Integration Theory to Explore the Associations Between Users’ Exercise Motivations and Fitness Technology Feature Set Use1. MIS quarterly. 2019 Mar 1;43(1):287–312.

9. Khurana R, Goel M, Lyons K. Detachable smartwatch: More than a wearable. Proceedings of the ACM on Interactive, Mobile, Wearable and Ubiquitous Technologies. 2019 Jun 21;3(2):1–4.

10. Till C. Exercise as labour: Quantified self and the transformation of exercise into labour. Societies. 2014 Aug 28;4(3):446–62.

11. Swan M. The quantified self: Fundamental disruption in big data science and biological discovery. Big data. 2013 Jun 1;1(2):85–99.

12. Hendrix S. Switching Wearable Fitness Devices: Exploring Factors in Switching. Madison (SD): Dakota State University. 2025. Available from: https://scholar.dsu.edu/theses/486/

13. Soliman W, Rinta-Kahila T. Toward a refined conceptualization of IS discontinuance: Reflection on the past and a way forward. Information & Management. 2020 Mar 1;57(2):103167.

14. Jamwal M, Kanojia H, Dhiman N. What motivates users to continually use wearable medical devices? Evidence from a developing nation. International Journal of Pharmaceutical and Healthcare Marketing. 2024 Feb 15;18(1):47–66.

15. Clawson J, Pater JA, Miller AD, Mynatt ED, Mamykina L. No longer wearing: investigating the abandonment of personal health-tracking technologies on craigslist. InProceedings of the 2015 ACM international joint conference on pervasive and ubiquitous computing 2015 Sep 7 (pp. 647–658).

16. Jeong H, Kim H, Kim R, Lee U, Jeong Y. Smartwatch wearing behavior analysis: a longitudinal study. Proceedings of the ACM on interactive, mobile, wearable and ubiquitous technologies. 2017 Sep 11;1(3):1–31.

17. Epstein DA, Caraway M, Johnston C, Ping A, Fogarty J, Munson SA. Beyond abandonment to next steps: understanding and designing for life after personal informatics tool use. InProceedings of the 2016 CHI conference on human factors in computing systems 2016 May 7 (pp. 1109–1113).

18. Esmonde K. ‘There’s only so much data you can handle in your life’: accommodating and resisting self-surveillance in women’s running and fitness tracking practices. Qualitative Research in Sport, Exercise and Health. 2020 Jan 1;12(1):76–90.

19. Tapia SM. Running Naked: A Thematic Analysis of the Lived Experience Running with and Without Technology. California Institute of Integral Studies; 2023.

20. Clark M, Southerton C, Driller M. Digital self-tracking, habits and the myth of discontinuance: It doesn’t just ‘stop’. New Media & Society. 2024 Apr;26(4):2168–88.

21. Mertala P, Palsa L. Running free: recreational runners’ reasons for non-use of digital sports technology. Sport in Society. 2024 Mar 3;27(3):329–45.

22. Clarke V, Braun V. Thematic analysis. The journal of positive psychology. 2017 May 4;12(3):297–8.

23. Malterud K, Siersma VD, Guassora AD. Sample size in qualitative interview studies: guided by information power. Qualitative health research. 2016 Nov;26(13):1753–60.

24. Tracy SJ, Hinrichs MM. Big tent criteria for qualitative quality. The international encyclopedia of communication research methods. 2017 Apr 24:1–0.

25. Braun V, Clarke V, Hayfield N. ‘A starting point for your journey, not a map’: Nikki Hayfield in conversation with Virginia Braun and Victoria Clarke about thematic analysis. Qualitative research in psychology. 2022 Apr 3;19(2):424–45.

26. Strava. (n.d.) Strava [Mobile app]. Apple App Store. https://strava.com

27. Lima CS. The Impact of Gamification in Smartwatches on Goal Pursuit and Engagement: Mediating Effects of Addiction and Self-Identity (Master’s thesis, Universidade NOVA de Lisboa (Portugal)).

28. Chotpitayasunondh V, Douglas KM. The effects of “phubbing” on social interaction. Journal of applied social psychology. 2018 Jun;48(6):304–16.

29. Garrido EC, Issa T, Esteban PG, Delgado SC. A descriptive literature review of phubbing behaviors. Heliyon. 2021 May 1;7(5).

30. Aslanova MS, Valieva AS, Bogacheva NV, Skupova AM. Mobile Food Tracking Apps: Do They Provoke Disordered Eating Behavior? Results of a Longitudinal Study. Psychology in Russia: State of the art. 2024;17(1):67–83.

31. Siddika A, Ellithorpe ME. I did 10,000 steps so I earned this treat: problematic smartwatch use and exercise tracking associations with compensatory eating and sedentary activity. Cyberpsychology, Behavior, and Social Networking. 2025 Mar 1;28(3):211–6.

32. Department of Health and Social Care. No health without mental health: A cross-government mental health outcomes strategy for people of all age. The Stationery Office. 2011. Available from: https://assets.publishing.service.gov.uk/media/5a7c348ae5274a25a914129d/dh_124058.pdf

33. Department of Culture, Media and Sport. Capturing engagement numbers: Stand 2 participation survey data analysis. Gov.uk. 2026. Available from: https://www.gov.uk/government/publications/capturing-engagement-numbers-strand-2-survey-data-analysis/capturing-engagement-numbers-strand-2-participation-survey-data-analysis

34. Bamidele OO, E. McGarvey H, Lagan BM, Chinegwundoh F, Ali N, McCaughan E. “Hard to reach, but not out of reach”: Barriers and facilitators to recruiting Black African and Black Caribbean men with prostate cancer and their partners into qualitative research. European Journal of Cancer Care. 2019 Mar;28(2):e12977.

35. Department for Science, Innovation and Technology. Online Safety Act: Explainer. Gov.uk. 2025. Available from: https://www.gov.uk/government/publications/online-safety-act-explainer/online-safety-act-explainer

36. Ummels D, Beekman E, Braun SM, Beurskens AJ. Using an activity tracker in healthcare: experiences of healthcare professionals and patients. International journal of environmental research and public health. 2021 May 12;18(10):5147.

37. Smith AA, Li R, Tse ZT. Reshaping healthcare with wearable biosensors. Scientific Reports. 2023 Mar 27;13(1):4998.

