## Supplementary Table 1 for "*“I had to learn to trust my body again”:* Exploring the emotional and behavioural impact of wearable activity tracker discontinuation and reasons for removal"

Drives for device discontinuance

| Data fixation | *‘I don't want to admit how much I did really enjoy it, but I did’ (P1).*  *‘I really did use it a lot, so I would track all my activity on it. But then I would look a lot at, you know, like how many steps I'm doing. Have I closed my rings and so like, have I exercised, how many calories I've burned, things like that, not to like, stick to a strict diet, but I just. The data was there so I couldn't help but look at it, which sounds like I have no self-control, but actually when it's all in front of you to like such detail, it's hard not to look at it so. I looked at everything’ (P1).*  *‘I actually really loved my watch, but that was the issue, I just got so into it’ (P1).*  *‘I wanted to stop using it all the time 'cause it was getting a bit obsessive. That's probably the closest words you'll get. I feel like I was scrambling for feedback the second it was available like when I finished tracking a run’ (P2).*  *‘I felt I cared about the data more than like the day out’ (P2).*  *‘I've also tried tracking it but not looking at my watch and I just I just couldn't really do it if it's there and on I do look at it, so, yeah, I still think taking off is the best option’ (P2).*  *‘That dependence on on my watch to tell me if I was getting better and tell me how I'm doing essentially was the issue’ (P2).*  *‘That's why I stopped using a watch on that one run; I was dependent on my watch’ (P2).*  *‘I think I was looking at the numbers way more than I would have preferred to normally’ (P3).*  *‘I was dependent on it before, I looked at it all the time’ (P4).*  *‘I didn't really have much else on. I was all in on the rings and then that stayed for like a couple of years. That was like a fixation’ (P5).*  *‘I was wearing it every day. And I was still very much into like the rings, and like, even though sometimes I would reduce the rings, so I hit it, but still I had to close the ring. At one point I had like, it was like a 1000 something day streak of closing my rings. So that will tell you how long I used it for. The issue is, is when he gets to that point of the streak as well, it's full commitment. The pressure builds’ (P5).*  *‘In the past, I would be like oh my god, I don't have my watch on, it's the worst thing in the world, the steps don't count… there'd be times where before I'd be like, I'm going to wait half an hour before I go out so that my watch can be charged so I can track my steps’ (P6).*  *‘It slowly consumed my life’ (P7).*  *‘It was the all consuming, like I was really consumed with the metrics that it was giving me, and no matter how arbitrary they were, I was concerned with my sleep score and I was spending more time really concerned with the aspects of like hitting my step goal, hitting my caloric intake, hitting my sleep goal, hitting all those metrics, no matter how good they were for me, I was more concerned with the number than the actual like benefit that they had for my life’ (P7).*  *‘It pulled a thought that existed in my brain, but I hadn't really explored, that was maybe I am using this a bit too much, specifically like looking at it too much because I was always checking my watch, you know. I would check it all the time and I didn't get notifications sent through to my Garmin. I think people will have presumed I'd got a text or an e-mail had been sent through, but I turned that off, so I was checking my watch screen. It could be every couple of minutes on a walk to see what my step counts gone up to and things like that’ (P8).*  *‘It started feeling like I was doing things to please the watch. I was like, you know, oh, the watch is telling me today I haven't walked enough, so I need to walk more and stuff like that, and that's not how it should be… so when I started noticing that in me, first I tried to regulate it and then I was like, okay, I failed to regulate it, so take it off’ (P9).*  *‘Four months after, and I got addicted already in four months to the beast’ (P9).*  *‘When it progressed to an Apple Watch and it got a lot more, I would say, obsessive maybe is the correct word’ (P11).*  *‘I think that's when I kind of realised that there was a lot of unhealthy habits going on and I didn't want my Apple Watch to control what I was doing in the day or anything like that. And if we'd gone out or something and I knew that we were going out the next day at uni, I would tend to get up and like go for a walk around the city centre just to get my steps in for no other reason. It wasn't to help me or anything’ (P11).*  *‘I realised there was much bigger things going on and I am obsessed about a little Apple Watch and what food I'm eating every single day…It feels a bit like a jail. Like I was in an Apple Watch jail’ (P11).*  *‘The first thing I would do when I woke up is like check the Fit app and like make sure that I had a good sleep. And I was like, I'm not paying any attention to how I actually feel in myself or in my body. Like I'm relying on an app to tell me whether I like had a good sleep or not’ (P12).*  *‘I was using it for everything. Like really, really using it a lot, tracking everything. But it just got a bit futuristic and I got a bit too into it. I was just, I was just paying more attention to it than life, essentially. And I did, I really enjoyed it. I really looked at the numbers. I saw every day as like a bit of a competition. But that meant I was turning down social plans like going to watch a film or, you know, um I don't want to do this because it's going to be 4 hours of driving in total and that's a big chunk of my day. Weird working out logistics like that and deciding if to do something almost based on the window I had left to keep beating my scores’ (P13).*  *‘That use all the time is what led me to just kind of forgetting the enjoyments of life and movement and the connection’ (P13).*  *‘I had started using it for loads of things and probably putting it over my brain quite a lot… I was just putting it over my brain and that really didn't sit right with me’ (P14).*  *‘You get into a routine and it becomes very structured and objective. But I think I was applying that same logic to moving and to my running, but also to like everything was, oh, can I get a little bit more in there? You know, I think some days I should have just driven to a shop and I was walking just because it's a bonus of steps, which is really good, but it comes to it comes to a point of, come on now, you're wasting time’ (P14).*  *‘I took it off because I was glued to the data. Like, you said, how do you feel about the data? And I thought, yes! I was looking at those numbers a lot, really too much’ (P14).*  *‘I got into a bit of a cycle of like pushing, pushing, pushing, trying to work out like every day and then falling off that bandwagon and stopping. And I felt like it was some weeks I was glued to the numbers and really invested and others I wasn't’ (P15).*  *‘I got diagnosed with PMDD a couple of years ago now and it was linked. I tend to want a lot more food and have like real mood swings and feel terrible about myself, like it's linked to my cycle. So I think sometimes I'm absolutely fine, but it's when I'm already vulnerable, when I'm due on and when I'm ovulating that I, yeah, I just got down, I got stressed, I got a little bit like, all or nothing and numbers focused. It could have been the best workout ever, but if I wasn’t pleased with the numbers it’s all I cared about’ (P15).* |
| --- | --- |
| Data concerns | *‘It's interesting, trail running as well, because I'm not running around a track. You know, you could actually do your best effort on a bang average mile, and so it's not your quickest, but because of the terrain, maybe there's loads of rocks and like footwork needs to be very technical that's going to slow you down or there's uphill sections or it's flat, but it's a complete bog. I think the fact that I do my long run specifically like in the trails, it also highlighted that I was trying to use a watch to judge my performance. When the watch can't actually judge your performance. It's looking at speed and distance, and in trail running it's not just speed and distance. It's like so much more than that. It's the weather, it's the wind direction, it's the footwork you need to do. So it was insight that the data isn't accurate in these specific conditions, but then also the understanding that I was treating it as accurate. I was treating it as gospel’ (P2).*  *‘I used it for everything and I also found it get me... I get frustrated at any like lagging. I had good watches, but they still, you know, you didn't have signal. This is slightly inaccurate. They were lagging and I, so I used it so much. I think I noticed its flaws’ (P4).*  *‘The numbers don't really like correlate with anything. Like steps, yes, that correlates with how many steps you've taken, but like how it knows exactly how many calories I've burned is so like, it doesn't, it can't know that because there's passive calories, there's active calories, there's so many things like that. And how it knew my sleep score, I don't even know what the sleep score really measured because I don't even know, it was just a number that it gave me’ (P7).*  *‘I used to try to justify it by like, oh, well, I'm just one person, but I don't like having to justify the data that's being collected on me’ (P7).*  *‘I know that there was a big deal in the United States, especially in states where those access aren't a right anymore, like they had to delete period tracking apps because it knows your geographic data, it knows when you're tracking your cycle, so it could tell if you got pregnant. And then it would tell if like you travelled to a state and then you could get in trouble because it knew where you were going. It was a crazy thing for a while. And so period tracking apps are not it. Oura has that built in and I didn't love that aspect of it’ (P7).*  *‘There's no way to opt out of the temperature tracking in Oura either, it just does it automatically and assumes when your period is based off of the temperature that your body is’ (P7).*  *‘I've also learned that it's only approximation. It doesn't really count the steps. It's not really accurate in very many dimensions. So you take it a bit more with a pinch of salt. It gives you the indication pretty much, you know the trend’ (P9).*  *‘I understand where other people are coming from, but for me it's, I also don't trust the accuracy of the software because I do stuff like, you know, heart rate training and then I'm like, I am definitely not in zone 4. I'm in zone 5 because I'm working so hard. And then if I try to keep to zone 2, it's like I'm barely walking because it, yeah, so it's not accurate. So I don't really listen to it. If it says, oh, your health is whatever, whatever metrics, it's, yeah, you need to go to an actual, actual, I don't know, lab to get your VO2 max and all that. So it's all not that accurate, so I don't really care too much about it’ (P10).*  *‘Even my max heart rate is like, I like, I think the first week I was going beyond the max heart rate, like my, the heart, the measured heart rate is beyond the max heart rate. So I was like, clearly it's not very good at calibrating’ (P10).*  *‘I come back to the accuracy a lot and I just think about, for example, in a race the time's usually the same because you start and finish at a different time, right? It's the same, but the distance is always slightly off. Same for things like heart rate. Some of the people at the club, some of my friends, they only do heart rate training. But for me, my heart rate varies so much depending on, I don't know, if I've had a drink before, if it's been someone's birthday, if I've got an injury and I'm having to work harder, and I don't know if that's just me, but it just feels a bit a bit wishy-washy’ (P14).* |
| Escaping unwanted data | *‘I started a new job that was more more demanding, like mentally, but also I was used to doing .7 like full time and this was full time so it was also more time and I also got quite I remember when I went to take it off I'd been quite poorly a couple of times and it was it was just a reminder of that I didn't feel good. And it was like nagging me, and I felt like I was teasing myself with this data as well, like, again, if it's in front of me, I will look there so I basically decided while I was still ill, to take it off’ (P1).*  *‘The end of it was I felt like I was being reminded that I wasn't great, and then it was making my mood worse. And it was a bit of a cycle’ (P1).*  *‘I found myself really beating myself up when I got busier with a job and and when when I got ill and… a watch telling me to move when I couldn't was the last thing I needed. It genuinely made me angry and it got to the point of me questioning. Hang on a minute, why is this on my wrist still?’ (P1).*  *‘I did used to get like an e-mail summary each week. Yeah, which is often how I like based a lot of how I felt about the week or what aims I wanted to have for the following week and I think like that was interesting but it also was one of the things which used to like affect my feelings about how my week had been’ (P3).*  *‘My watch didn't recognise that and all of a sudden it went from these nice messages of what I perceived as encouragement to like heckling. And it was, it was the same message, really. It was, it was just the move notifications. It was like if I did meet a goal, it was late, but then on my phone it was telling me that everything was down as well. The sleep scores I got in the morning were bad and I thought, hang on a minute, it's only going to get worse. It became negative and it was this weird, like, I do care, but not right now’ (P4).*  *‘It's not telling you anything you don't know. I know I'm tired, I know I'm worried, I know I'm stressed, I know I'm not working out as much as I was before. The only thing it's bringing me is like feeling bad about it, so I took it off’ (P4).*  *‘The watch before baby made me feel great. The watch after baby and during baby made me just feel bad about what I was doing’ (P4).*  *‘I used to like work two jobs and was a master's student and then I started full-time work. Like of course my steps went down, but then just makes you feel worse because it's like, oh crap, the red line's lower than it was over the last three months period or whatever’ (P5).*  *‘It finds a way to like feed into the doom scrolling aspect of like having a screen. Like it finds a way to really hit on those dopamine like indicators. And I just, it gave me an icky feeling after a while’ (P7).*  *‘We get these relapses, you know, so we get moments in which, well, I do get fatigue. Where you know, so it's not very nice. It's not really helpful because it does nothing but remind me of the condition that I'm in, you know. So it doesn't really help’ (P9).*  *‘I don't need to know that much data. Yeah, like if I, if I don't get good sleep, I know I don't get good sleep. I don't need my watch to tell me, and sometimes it's like, yeah, just additional data that isn't useful to my life’ (P10).*  *‘I do think it's unnecessary for me because I'm, yeah, that's an additional thing for me to worry about. I'm like, I have enough things to worry about. I don't need my watch to tell me that I'm not sleeping well. I know’ (P10).*  *‘I'm a normal person. I'm not looking at data in the morning going through, oh, how many percent REM sleep? How many percent stage 1 sleep? I don't think that's useful information’ (P10).*  *‘I'd never wear it to the office and that is because one, the reminder to stand, that would annoy me, and then two, would probably be because of how you can see your message and stuff like that. And I'm a one that if I go on my phone then I'm not getting any work done’ (P11).*  *‘I got injured and I was still feeling this injury and like January, February, March. So what happens at this age, the injuries last longer. And then because I had a lot of time to rest, I could feel my shin as well. It was tightening and I thought, I've never had shin splints before and as if it's going to be now and I managed to stave it off, but I wasn’t running and I just didn't didn't like the data because I already had such a hard time stopping, not from this like compulsive level, but because if you get into a sport like this for so long, it's because it's good for your mind. You're doing it for a reason. There's lots of benefits. I had to stop for a bit and the watch was just reminding me I hadn't run for a while when I was stopping, but then when I put it back on... I was comparing everything to my previous fitness and you do lose fitness quite quickly in running. It wasn't, the watch wasn't saying, oh, this was worse than six months ago. It wasn't directly saying that. However I'd become so used to the numbers. I knew my numbers. I knew how I was doing this on scores. So I could see the scores had changed’ (P14).*  *‘I was a bit down that I knew when I was doing this good thing. I was just thinking, oh, I'm not as good as before. Of course you're not, you were injured. But it was just this awareness. That made me feel a bit like shit, even during running these huge distances. That reminder this number's gone down, this number's gone down, this number's gone up. It wasn't nice. It was distracting me from the actual doing and the seeing and making me forget’ (P14).*  *‘I just got in a bit of a bit of a state and I thought well let me try taking it off because I was noticing I'd feel bad for not moving some days, and my job is remote, so you know some days I really don't move, and it was, I wasn't glued to it, I don't look at it often but it was still sending me like notifications to move and I just felt like it wasn't helping my brain’ (P15).*  *‘It wasn't this huge pressure to completely change, but if you were debating one or the other, you'd go for the better option with the watch on…Just aware of some kind of responsibility. Which sometimes I loved but other times made me feel a bit itchy’ (P15).* |
| No longer aligned with lifestyle or values | *‘I started a new job that was more more demanding, like mentally, but also I was used to doing .7 like full time and this was full time so it was also more time and I also got quite I remember when I went to take it off I'd been quite poorly a couple of times and it was it was just a reminder of that I I didn't feel good. And it was like nagging me, and I felt like I was teasing myself with this data as well, like, again, if it's in front of me, I will look there so I basically decided while I was still ill, to take it off’ (P1).*  *‘I found myself really beating myself up when I got busier with a job and and when when I got ill and… a watch telling me to move when I couldn't was the last thing I needed. It genuinely made me angry and it got to the point of me questioning. Hang on a minute, why is this on my wrist still?’ (P1).*  *‘I've started trying to have like a slower life, life at a slower pace, and like more relaxing, and I think taking it off is something that's really helped me’ (P1).*  *‘One of the reasons I took it off was because I just got a bit into this idea of the fact we need to improve at all times and I was definitely just tired and on the road to burn out from this watch’ (P1).*  *‘I think the time I had off from it, I think I was also just in like a space online and things like that, which was kind of being about not like having to exercise. You don't have to force yourself to do it for a specific body type, a body positivity kind of like lens that I think I grew into a lot more during that time where I wasn't wearing it’ (P3).*  *‘I don't think it's very natural, like unless like you have to have a machine and things tell you that you're doing a good job. I think you should be able to find that within yourself’ (P3).*  *‘My wife had a baby and she was extremely unwell, so my priority went from being the fittest I could be to helping my wife get through the day’ (P4).*  *‘Exercise, no longer did it become my priority to get this many minutes a week of this and this, don't get me wrong, would it have still made me feel good? Yes. Did I want to do it? Yes, but life happens and it just shifts your perspectives of what's important. So my friend had already planted the seed. My wife was just having the worst pregnancy. It was truly rough and it was a long time if you're having a bad pregnancy. So my priority changed there, and stating the obvious, my watch didn't recognise that and all of a sudden it went from these nice messages of what I perceived as encouragement to like heckling. And it was, it was the same message, really. It was, it was just the move notifications. It was like if I did meet a goal, it was late, but then on my phone it was telling me that everything was down as well. The sleep scores I got in the morning were bad and I thought, hang on a minute, it's only going to get worse. It became negative and it was this weird, like, I do care, but not right now’ (P4).*  *‘I thought I would return to it but my sleep's not great now. Spoiler alert - with children, sleep is bad. It's just like I'm living a different lifestyle now. I still like hiking, I still like swimming. I've slowly picked up all of the activities, but it's can you fit this into your busy life instead of I need to do this on this day to like get this score’ (P4).*  *‘The watch before baby made me feel great. The watch after baby and during baby made me just feel bad about what I was doing’ (P4).*  *‘I started work, obviously my like activity levels like went off a cliff because it's like a full time, like very long hours job’ (P5).*  *‘I think part of the reason that I, I mean, part of the reason I bought this because, you know, like it was in lockdown, I was trying to get activity in, I was trying to lose weight, all of this. So it's always been kind of prompted by like concerns about like my weight and my appearance. And I think I've also been trying to make like a more intentional shift to like doing activities that I enjoy when I want to, and I think that the watch doesn't really fit into that vision’ (P5).*  *‘At the moment I'm trying to just be a bit more mindful about like my approach and doing things when I like where I enjoy them and stuff and yeah I just don't think that the watch helps me with that?’ (P5).*  *‘I think me trying to like be a bit easier on myself kind of led to not wearing the watch as much’ (P5).*  *‘I used to like work two jobs and was a master's student and then I started full-time work. Like of course my steps went down, but then just makes you feel worse because it's like, oh crap, the red line's lower than it was over the last three months period or whatever’ (P5).*  *‘My life has changed, like things that were reasonable, no longer reasonable, but that doesn't have like some, you know, like, good or bad, like moral bearing on like my body or me’ (P5).*  *‘I was putting pressure on myself, to, you know, like go for a walk, go to the gym. I'm not doing that. It's the 23rd of December, but I just, I just felt this like avoidance and hesitation and it was, it was just getting up to Christmas and we've got large families so we do a lot, like we try and see everyone. So actually we're probably active, but we have no flexibility and no free time. It's just, it's a lot of sitting on sofas chatting, like once you get to the place. And it got to a couple of days into when I'd had time off work, that stretch, and I needed to charge it and I just took it off to charge and you know it's so quick charging but something just like clicked and thought I'll just put it back on in like January. I'll just put it back on after Christmas. I hadn't decided this like set date but I just thought if it's getting in the way now you're going to be doing even less and indulging more in a week’ (P8).*  *‘In the back of my mind I felt like it was gonna tell me off that enjoying Christmas, essentially, so I wasn't dependent on the data, but I still saw the data was really important. I obviously cared about the data’ (P8).*  *‘I started getting annoyed by the buzzing because, you know, you fall back in your old habits and stuff and you're like, come on, I'm doing something. I can't move now, you know, that sort of thing, and then I started realising that I was like too much under its control, I don't like to be controlled’ (P9).*  *‘I don't compare my fitness anymore as a statistic. And I was, I used to, it was purely about how fast I could do a 2K run or how many calories can I burn in a HIIT workout. Like it was kind of, I just thought about it in the numbers. It was never about like my own actual health and fitness’ (P11).*  *‘I'm just not really, the point right now, I'm not really bothered by it. Whereas if you would have asked me four years ago, I would have probably died on the hill for my Apple Watch. It was, it was honestly meant everything to me. Like if it, the most old destroying thing is if you, if I got up, put it on and then was carrying about my day and it got to like 10am and it died, like I just feel like my day is wasted. Like I could have just gone home and gone to bed at that point because what's the point? What is the point of my day if I can't like track everything on my Apple Watch? But yeah, not like that anymore’ (P11).*  *‘I just felt that, yeah, I was paying too much attention to an app rather than my own body and so it just didn't feel very like genuine’ (P12).*  *‘Okay, you've had this one wake up on your health, a wake up call on your health, sorry, but what's the next wake up call going to be? Are you going to have missed your last time to see somebody because you were going on a walk, but not even a walk you were really wanted to do’ (P13).*  *‘I had started using it for loads of things and probably putting it over my brain quite a lot… I was just putting it over my brain and that really didn't sit right with me. I pride myself in not being dependent on technology because so many people are, you know, especially these younger generations, I see them glued to their phone and it does upset me. I think you're not looking up and I don't know if you're talking to your friends, they're in a huddle and they're all looking down. And I almost had a bit of a, not instant, but a recognition over time from looking at other people on these runs, but then thinking about what I do alone and realising I'm kind of doing that with a watch. That's my focus. I'm not getting anything else out of it, out of running, because I'm staring at it. So it just didn't sit right with me as well in terms of who I think I am as a person, but then what I wasn't like following through with’ (P14).* |
| Others approval | *‘It was actually a friend. I was pretty influenced by them, but I was getting frustrated and I was chatting to them and they've always been very, I don't think they've ever used a smart watch, They like do everything just with their Casio on, which stands out from the crowd nowadays and I'm pretty sure it came about because we were working out together and he made a comment on, like you essentially won't stop looking at your watch but what's it going to tell you that you don't know if you’re doing it right here?’ (P4).*  *‘It was another time after that he said, seriously, I don't think I've seen you look at that watch and look happy from whatever it's telling you. And so I didn't take it off straight away, but he definitely planted the seed’ (P4).*  *‘One of my friends said to me at one point, she was like, this is absurd, like, why do you have such a long streak on your watch? And I was like, that's such a good point’ (P5).*  *‘My wife asked me about it because I said that I was going to do this. She joked and was like, it's like you're not checking in with your boss like every day. It was like it was like a micromanager’ (P8).*  *‘I didn't realise about the whole like, you know, when girls wear Apple watches to go out and just like take it off kind of vibe, like all them memes going around and people would say, like making fun of that basically. I didn't know that was going on, I didn't know that was a thing. So then when I came to university and we were going out with freshers, everyone was like, not laughing at me, but it was part of the punchline that I was always wearing my Apple watch’ (P11).*  *‘I did have one friend who, she got an Apple Watch when we started uni, but she got a bit obsessive too and then kind of went away from it. And we do talk about it quite a bit, just like tracking things and we've all come to the conclusion that it's not good for any of us. So we just don't do it, really. And I think a lot of people that I know are like that’ (P11).*  *‘[My mum] hated me with it, like really didn't like me with it… my mum was over the moon. She had been trying to get me to do it for a long time, so I think that was good for her’ (P11).*  *‘I think my wife had been noticing for a while and, yeah, after a while she mentioned it and I was in, I was properly in denial. But she was saying, you know, you're almost, you're choosing to have more time to move over, seeing your kids and seeing some of your best friends’ (P13).*  *‘I thought, do you know what? I think I'll be exactly the same, but I'll take it off, not just to prove a point, because annoyingly I've come to know after decades of marriage, my wife is often right… I thought at the time, well I don't think it's an issue. I'll listen to her, but it'll backfire. It won't be any different’ (P13).*  *‘I'll see people tracking things and every so often without sounding stupid, I do feel a little bit envious. I do look and I think, oh, I won't want your having, that's nice. But family comes first. Family absolutely comes first in terms of, yeah, just life’ (P13).* |
| Purpose of activity | *‘I do a long run every Saturday or Sunday, that's over two hours, and it's that run where I'm not focused on, you know, running intervals at a certain pace that it's not the run that's designed in my plan to have really strict thresholds I need to meet. It's the run to go and do just to build endurance. It's about getting out there, so I made a bit of a pact maybe like four months ago now for my long runs to just be without a watch, just go out and do it’ (P2).*  *‘The whole point of these runs is distance, not speed, but I was still looking at speed and I was still having this that made-up speed in my mind that I wanted’ (P2).*  *‘I found like at the start of the class, I was like sitting on my watch, like scrolling through, trying to find why for yoga to add it on. And like, it was just like taken away from like the chill sort of calm experience. And again, yoga is not something that I worry about where I'm like, I'm not super doing yoga for exercise. It's more for like my mental health and like to relax me and to calm down so I don't want to be like, oh, how many active minutes have I had?’ (P6).*  *‘I just feel like in yoga, I don't worry as much about like, what's my heart rate going to be just now? Or am I expending enough energy like doing this exercise? I just don't think about it whatsoever and it's really nice to not second guess or try to predict like, oh, the watch is going to say X amount of calories’ (P6).*  *‘I definitely wouldn't choose to wear it if I knew I was having a day where I was sitting down a lot because that would then make me think about it and I wouldn't want to think about it’ (P11).*  *‘I'd look at data at the end and be like, oh, it felt much harder and I'd kind of tear apart what I'd done. I would be like, what, it felt harder than that or maybe this wasn't good for me, when actually I’m kind of forgetting the whole point of doing it, of moving, of feeling good’ (P15).* |
| Forced discontinuance | *‘I think initially I stopped using it because it broke. And so like the strap broke and I had to replace it. So it was one of those things that I actually just like couldn't wear it, and then I procrastinated getting a new kind of strap, and then I think I'd gotten a new one, but by that point I was kind of just out of the habit of wearing it’ (P3).*  *‘I think like the strap broke again or I think I just wasn't enjoying wearing it quite as much so I put it down and yeah left it’ (P3).*  *‘The battery life on it isn't very great because it's an Apple Watch, so like if it's dead and then I need to wait like 15 minutes for it to charge, I'm like, I can't be bothered, and that's how it sort of started’ (P6).* |
| Device or platform switching | *‘I quite like having the latest and greatest for no real reason other than I. I like, I do like upgrading things’ (P8).*  *‘I decided I want to see what it was like with a screen, and because I was looking at a screen for so long, I thought, how would it be if there's no screen?’ (P8).*  *‘I downloaded Strava, so I could get kind of that more in-depth detail in terms of like pace and like distance, so I could plan a route and stuff like that, stuff that I could do and then I realised that I was relying on Strava more and I wasn't really interested in kind of what the Fitbit was telling me. Like I prefer, I would rather go based on how I feel than how a watch tells when a watch kind of tells me to recover or when a watch tells me I've not enough sleep or something. So eventually I just took it off’ (P12).* |
