## Supplementary Table 2 for "*“I had to learn to trust my body again”:* Exploring the emotional and behavioural impact of wearable activity tracker discontinuation and reasons for removal"

| Theme | Sub-theme | Quotes |
| --- | --- | --- |
| Attitude change to activity | | |
|  | Judging by feeling, not numbers  (QS) | *‘I would try and look at my screen and see oh I only burnt this many calories so it can’t be useful’ (P1).*  *‘I was trying to use a watch to judge my performance. When the watch can't actually judge your performance. It's looking at speed and distance, and in trail running. It's not. It's not just speed and distance. It's like so much more than that. It's the it's the weather, it's the wind direction. It's the footwork you need to do so it was insight that the data isn't accurate in these specific conditions, but then also the understanding that I was treating it as accurate. I was treating it as gospel’ (P2).*  *‘It's having a really nice run, not particularly nice in terms of things like weather because it's horrible at the moment and really brutally windy, but I've had like, a rewarding run. I've just got out and instead of, you know, saying that was good, that was rewarding, I then look at data straight away and feel a bit disappointed. So it was, it was taking away from the fact that I was doing something good and the whole point of these runs is distance, not speed, but I was still looking at speed and I was still having this that made-up speed in my mind that I wanted’ (P2).*  *‘I think in the moment I'm calmer or maybe not specifically calmer, but if I get to a section that you know would typically slow me down, perhaps. There's a big puddle like we get bogs up here on the malls like real bogs. And I'm not thinking, oh, for God's sake, that's going to slow me down. I'm just like, yeah, I'm on a hill. I'm going for a bog. It removes that that annoyance there’ (P2).*  *‘I see my long run now as my like stress relief. It's really nice. I used to do them on a Saturday, every every weekend and I'm slowly moving them over to Sunday because I feel like it's just a really nice, it’s nice man. Like Sunday reset, we've got life admin and then you, you cook your lunches for the week and you go and wander and jump about on hills. And I feel like it's it's exactly what I need. It's not this like time trial’ (P2).*  *‘Now I'm having to judge what I'm doing from my body instead of my watch, which actually when you're used to looking at pace and speed and distance is very different. So I think even if I'm not loving it and I don't think it's like as in depth, I am having to listen to my body more on these long runs because the watch isn't telling me to speed up or slow down or the time I've been running for’ (P2).*  *‘I thought how will I know if I'm getting better if I'm not wearing my watch and that was the panic. But also, that dependence on on my watch to tell me if I was getting better and tell me how I'm doing essentially was the issue that was. That was why I was using data too much and why I was relying on it. So I'm almost having to make peace with the fact that I can't judge if I'm making progress on those long runs in that same objective way any more, but instead I can ask did I feel good? And I'm having to remind myself that generally you will be making some progress, you're getting outside, that's good for you’ (P2).*  *‘I have more recently come around to the idea of just like exercising like for health, but not tracking anything, seeing, just like doing it based upon like how you feel, your body, not necessarily focusing on numbers, anything like that. Calories burned, I don't really want to know all that stuff, so I think that helps me’ (P3).*  *‘I do it by kind of like metrics of what I've managed to like achieve in terms of like what weight I've managed to push or pull or left. But then also just kind of how I feel for the rest of the day after it or like the next day is like, am I really hungry after it? Like, is my body like craving sustenance because I've probably used it quite well or do my legs hurt the following day and things like that. And when they do that, it's almost like, oh, but like it feels good because you're like, okay, well, I, that means I'm changing my body’ (P3).*  *‘I don't think it's very natural, to have to have a machine and things tell you that you're doing a good job. I think you should be able to find that within yourself’ (P3).*  *‘We were working out together and he made a comment on, like... You, essentially, you won't stop looking at your watch but what's it going to tell you that you don't know if you’re doing it right here?’ (P4).*  *‘You know what you're doing, like you're doing it. You know more than a watch’ (P4).*  *‘I don't know now if I'm like 200 steps off getting my step count for the day, so I might naturally do that in the evening still, but I'm not purposefully going to go like up and down the stairs a couple of times to get that. So, yeah, sometimes I might do less, but I think it's better’ (P4).*  *‘It's just got to be how I feel now, which makes sense, really, doesn't it? It's not a bad thing, so yeah, I just see how I feel and that's all I can do’ (P4).*  *‘When I am on runs. I think previously I would think a lot about my pace and I would think a lot about my heart rate, which were both on the screen when I was running. And, you know, I would think, oh, you know, this pace is supposed to be like my easy pace. So why does it, you know, why does it feel hard today? Or like, I really need to stay within this heart rate zone. And I think I probably am running slower now. I'm not as fit as I was, but equally, I go out for a run and I'm like, hmm, I feel like I'm running a bit too fast, like I feel a bit too out of breath, so I'll slow down and I definitely think it's just helps me do things like at an easier pace rather than saying like, okay, this heart rate and this pace is easy and so I have to stick to that’ (P5).*  *‘Enjoyment is just much more about, like, I focus a lot more on how I feel physically. Like, you know, sometimes I'd be running with my watch and I'd be like, okay, I'm in my easy pace range, but I'd literally be like gasping for breath. Yeah, whereas now, it's much more, like, you know, physically, like, does my chest feel tight? Like, how do I feel? You know, how do my, like, my ankles and my hips, like, feel today?’ (P5).*  *‘I'm not super doing yoga for exercise. It's more for like my mental health and like to relax me and to calm down. So I don't want to be like, oh, how many active minutes have I had?’ (P6).*  *‘I'll go, oh, that was like a good workout because I burned like X amount of calories and that's not really a mindset that I want to be in’ (P6).*  *‘I think like the removal of it has really helped me to like reframe like, oh, as long as I'm moving, that's all that matters. Like I don't need to do X amount of hours a day as long as I'm moving a little bit, like a little bit is better than nothing. And I think it was easier for me to not have that absolute or all or nothing feeling towards like exercise and sleep and things like that’ (P7).*  *‘I think the only judgement is, is just those indicators that we were talking about. Like, do I feel at the end of the day, like I did enough? Do I feel, do I feel good in my body? Do I feel good in like my mental health and things like that? So, I think I don't have any like external metric. I'm not tracking like in a journal. I'm not tracking in anything like that. I'm just trying new things and seeing what sticks and then like feeling good about it or not good about it and then adjusting whatever I need’ (P7).*  *‘You don't have things like calories anymore, so you can't check that … the calories you burn from a full body strength training workout or a lower body compared to one just focusing on upper body, there is a difference and I know while I would have said I need to go to work out like I need to do it today, my calories are low, I've not done much. I would also find myself like shuffling it around and thinking, okay, let's do one of those workouts that is more intense when actually the others are pretty important, but I did start looking at calories burned’ (P8).*  *‘I don't need to know that much data. Yeah, like if I, if I don't get good sleep, I know I don't get good sleep. I don't need my watch to tell me, and sometimes it's like, yeah, just additional data that isn't useful to my life’ (P10).*  *‘I don't compare my fitness anymore as a statistic. And I was, I used to, it was purely about how fast I could do a 2K run or how many calories can I burn in a HIIT workout. Like it was kind of, I just thought about it in the numbers. It was never about like my own actual health and fitness’ (P11).*  *‘I probably wouldn't, I don't push myself as much anymore when I'm exercising because I would, that would be a bad habit of mine. Like I would go until I was throwing up because I just wanted to burn as many calories as possible. But now I know that that's not the purpose of exercise. The purpose of exercise is just go one for health and two for your own mindset and wellbeing, but I'll go because it feels good to move my body kind of vibe. But in the past it was it was purely for one purpose and that was just so it was it was for sure. It was I've burnt so many calories today. Like that was my whole purpose when I was, when I was constantly wearing it’ (P11).*  *‘I just don't really think about it in too much detail anymore. I think if I'm if I'm if I feel comfortable and happy, then that is that's enough for me now. Like, I don't need I don't need to measure it in any way, shape or form. Because I don't think, I don't, well, so I very rarely look at calories and things like that’ (P11).*  *‘The first thing I would do when I woke up is like check the Fitbit app and like make sure that I had a good sleep. And I was like, I'm not paying any attention to how I actually feel in myself or in my body. Like I'm relying on an app to tell me whether I like had a good sleep or not’ (P12).*  *‘I do see a lot of people working out in the gym now with one on and wonder are they in the same place that I was? Do they realise? You can't really ask without it being a bit of a like weird intervention, but I do think about it because it just becomes the normal behaviour. You just, you're properly using your watch, but that use all the time is what led me to just kind of forgetting the enjoyments of life and movement and the connection. It became nothing to do with how I felt, which is mad given it makes you feel so good. That’s what really matters. And yeah I just got a bit lost for a couple of years’ (P13).*  *‘I was comparing everything to my previous fitness and you do lose fitness quite quickly in running. It wasn't, the watch wasn't saying, oh, this was worse than six months ago. It wasn't directly saying that. However I'd become so used to the numbers. I knew my numbers. I knew how I was doing this on scores. So I could see the scores had changed’ (P14).*  *‘It was just this awareness. That made me feel a bit like shit, even during running these huge distances. That reminder this number's gone down, this number's gone down, this number's gone up. It wasn't nice. It was distracting me from the actual doing and the seeing and making me forget. You're going to run, yeah, because it's good for your fitness, but because you enjoy it’ (P14).*  *‘It was just when I was quite, like I felt vulnerable already and then it wasn't helping me. I have it a history of like, just a lot of like yo-yo dieting and things like that as well. And I got diagnosed with PMDD a couple of years ago now and it was linked. I tend to want a lot more food and have like real mood swings and feel terrible about myself, like it's linked to my cycle. So I think sometimes I'm absolutely fine, but it's when I'm already vulnerable, when I'm due on and when I'm ovulating that I, yeah, I just got down, I got stressed, I got a little bit like, all or nothing and numbers focused. It could have been the best workout ever, but if I wasn’t pleased with the numbers it’s all I cared about’ (P15).*  *‘It's not that I enjoy it more, but I, I don't dislike things as much after feedback. So sometimes I'd look at data at the end and be like, oh, it felt much harder and I'd kind of tear apart what I'd done. I would be like, what, it felt harder than that or maybe this wasn't good for me, when actually that was... I’m kind of forgetting the whole point of... of doing it, of moving, of feeling good. So I don't think I enjoy it in the moment more, but it's meant I stopped criticising what I've done afterwards and like downplaying it’ (P15).*  *‘It's just like, did I enjoy it? And like, was that hard? Did it feel, did it feel like a lot of effort? You know, some days you feel, you feel winded and you warm up and other days you feel strong… Just asking myself about it now, yeah, it's quite nice’ (P15).* |
|  | Attentional shift in activity | *‘I do a long run every Saturday or Sunday that's over two hours and it's that run where I'm not focused on, you know, running intervals at a certain pace, it's not the run that's designed in my plan to have really strict thresholds I need to meet. It's the run to go and do just to build endurance. It's about getting out there, so I made a bit of a pact maybe like four months ago now for my long runs to just be without a watch, just go out and do it’ (P2).*  *‘I felt I cared about the data more than like the day out’ (P2).*  *‘But I do think it's working quite well as a bit of a a reset. I think it's probably better for my mind. I feel like I was so focused on pace and speed when I used it on that long run. I wasn't actually getting the mental benefits of running. I'm getting physical because you're still moving, but it wasn't clearing my mind, it was focusing on something completely different, which was that data’ (P2).*  *‘If you run with a watch and do like pacing and heart rate work you get used to staring at your wrist for a lot of it. I mean, it's taken away from, like, just what you're doing, the way you're hopping along paths, the amount of views I must have missed or not fully taken in because I was looking at my wrist makes me pretty annoyed because, you know, I've done a lot of running in Scotland. I've done running in Ireland, I've done done some in Italy, like the amount of views I couldn't see, that you've missed from wearing my watch and being glued to it doesn't make me feel great’ (P2).*  *‘I said to my partner, it's like I'm just like, I'm just like a mountain goat and I'm just trotting around instead of before I feel like I was, I was trying to make things like too efficient. I was I was being really rigid, so I think it's helping with enjoyment. It's reminding me like why you run a bit, which I think you can get lost with if it's, if it's just speed, like, you're not just running to get faster, you're running because it's good for you and it makes you feel good and you enjoy it in the moment. And I honestly think I forgot that for a long time’ (P2).*  *‘I was in the mindset that you needed a watch to get better, to get fitter, you needed that data to train well and now it's changed my perspective’ (P2).*  *‘I think it's making me a little bit more balanced and chill. I still love it. I still worship the ground it walks on, I think it's just an amazing bit of kit, but I now see the place that running without a watch on can have and I do think it's so much better for my mind. It's making one run a week completely peaceful and just just time to talk to myself, time to reflect on my week, time to reset which I don't think I was getting before. I think before there wasn't that switch off because I don't know if you can fully switch off when you're looking at your watch that regularly’ (P2).*  *‘Having that data to be more competitive means you you can zoom in a bit too much and focus and think, oh, I just missed this. How did I not do that? Oh my, I felt a lot quicker on this stretch, but actually it was this. Whereas because I don't have that even though some days I'm really bitter that that is gone because I do enjoy looking at it, it makes me just appreciate what I've done without getting into the nitty gritty’ (P2).*  *‘I wouldn't want to know anything about like minutes I've been active or calories I've burned or like minutes I've been exercising. Like I don't think that would be as important to me’ (P3).*  *‘It's helped me be a little bit more relaxed about things, I think, and like possibly have a little bit more of like a long term view. I'm trying to like take a longer term view of like, it doesn't matter if in the next three months I'm not in the best shape of my life because I'm 26 years old. And you know, I've got hopefully like at least 70 more years on this earth’ (P5).*  *‘It's not progress over time, it's have you ticked everything off the to-do list? It's a very different, it's pressure to keep going instead of like this is what you've managed’ (P5).*  *‘I'm not like constantly checking my pace anymore’ (P5).*  *‘I've been doing yoga for like almost a year now. And I found like at the start of the class, I was like sitting on my watch, like scrolling through, trying to find why for yoga to add it on. And like, it was just like taken away from like the chill sort of calm experience’ (P6).*  *‘I just feel like in yoga, I don't worry as much about like, oh, how, like, what's my heart rate going to be just now? Or am I expending enough energy like doing this exercise? I just don't think about it whatsoever. And it's really nice to not be like, second guess or try to predict like, oh, the watch is going to say X amount of calories or heart rate, like, and especially in yoga , I think it's very good to not think about that at all’ (P6).*  *‘I was spending more time really concerned with the aspects of like hitting my step goal, hitting my caloric intake, hitting my sleep goal, hitting all those metrics, no matter how good they were for me, I was more concerned with the number than the actual like benefit that they had for my life’ (P7).*  *‘I'm a nicer person now. I was horrible. I was horrible to my mum. I was horrible to a lot of people around me because I was so obsessed over one thing. So I am actually a decent human being’ (P11).*  *‘It's not an urgent, like, let me check my watch and see how that exercise went. Like, like, I'll do it like days later. I'll be like, oh yeah, I did a class. And so it's not that initial sort of need for feedback or for like almost like acceptance from like the app or the watch to be like, okay, well, yeah, yeah, it feels like approval’ (P12).*  *‘I felt like supercharged. I just found this like cheat code to extra motivation and thought, where's this been my whole life? And I was using it for everything. Like really, really using it a lot, tracking everything. But it just got a bit futuristic and I got a bit too into it. I was just, I was just paying more attention to it than life, essentially’ (P13).*  *‘I miss having a watch, but I really, I do understand. I understand why. It's probably good as well in terms of I'd get really caught up on some numbers every so often. I would, I'd get really caught up and I'd almost fixate on my heart rate over the fact that, I don't know, I could have, I could have just got a new PB or I could have, you know, done an extra 2 reps on this weight that I've not done in a while. I would pick out one piece of data and focus on it, so I do think I'm just a bit more balanced. A watch could have told me one piece of information I wouldn't know otherwise that would make me forget to acknowledge the rest of the workout, so just a bit more zooming out’ (P13).*  *‘If my watch died during despite knowing that, I was dependent on the watch to like tell me I've done that, to verify it and in my mind it didn't count without it. So it's been a bit full circle and I can now see them for this really balanced view of absolutely incredible, but... maybe not something that should be permanent. Maybe it should be for educating people about the current movement, a current energy being burnt. And it sets you up with those good habits, but it's not an everyday life object’ (P13).*  *‘It was just this awareness. That made me feel a bit like shit, even during running these huge distances. That reminder this number's gone down, this number's gone down, this number's gone up. It wasn't nice. It was distracting me from the actual doing and the seeing and making me forget. You're going to run, yeah, because it's good for your fitness, but because you enjoy it…you're not pushing to represent the Olympics or something. It's more so you like doing it and it's good for you as a bonus. And I was, I think I was forgetting that a bit and I had just become a bit of a robot, I think’ (P14).*  *‘When I'm up on the hills, I'm taking in the run more. I'm noticing more, but I'm also noticing how I feel, because I'm not just looking at pace and heart rate. I'm thinking about it instead, because it's not there to beat my thoughts to it. Because it's so quick and it's constant you would look before you processed your own thoughts about it, really’ (P14).*  *‘I'm just more present, like looking at, for example, my run this weekend, quite a short one, but it was in an area I've not been to in a couple of months and I was just noticing all the daffodils and it was so cold, there was frost, but I was noticing daffodils. I was noticing signs of spring coming back and it was really nice and I know for a fact I wouldn't have noticed that and I did enjoy that. You're more connected with nature because you're not connected to just another computer, really. Let us not beat around the bush, it’s just a different screen. I think my enjoyment is the same, but before my enjoyment was from constant numbers, and now my enjoyment is from noticing what's going on, and, yeah, just looking around more’ (P14).*  *‘I'm simultaneously thinking about my body less in terms of like it just being all of these like numbers in and numbers out. But then I'm also thinking about the fact I'm thinking about it less. I don't know if that makes sense. I'm guessing this might calm down in a couple of more months because at the minute I've you know, it's still new to me. So at the moment, I'm like, wow, this is different. I'm now thinking about how it's different’ (P15).* |
|  | Regret over past behaviour | *‘The amount of views I couldn't see, that you've missed from wearing my watch and being glued to it doesn't make me feel great’ (P2).*  *‘I feel like having like removed it sometimes has given me a better relationship with this as opposed to my Fitbit when I would have it on every single day, you track every single calorie. If I had like 9000 steps, I'd be like, oh, I'm just going to go on a quick walk to get 10,000 in and it's like the 1000 is arbitrary at this point. So I think it has made me less. I don't want to say like dependent on the technology, because again, I'm on my phone, but like less dependent on the data and less bothered by it as well. If I don't reach whatever or however many steps, I'm just like, well [shrugs]’ (P6).*  *‘With 10 years of hindsight I can tell you that it probably wasn't good. I think, I just used it for everything and I got so into it. It wasn't necessarily terrible, but I really use it for tracking calories, for cooking. I basically let it determine what I did which seems odd to say now, but I do think 2016 was weird, and it was all, it was always normal then’ (P8).*  *‘If I looked at my steps for the day and say I hadn't done much, I just, because it was all online, all our lectures and things, so I wasn't really moving about a lot, and I hadn't joined a gym as of yet because I was kind of just settling in. So if I remember one day, I swear I did something like something ridiculous, like 400 steps and I actually cried, which was very embarrassing’ (P11).*  *‘I thought, oh my God, yes, it's in every single picture. That's really embarrassing’ (P11).*  *‘We'd been out the night before, me and my flatmates, and then I got up early to get my steps in and go for a walk, because at this point I was always wearing it. I hadn't wanted to go out because, you know, I can't do that anymore, get laughed at. So I put it, put it on, went on my walk and I was walking about trying to, I don't even know if I had a goal in mind. I think I was just trying to just go for what, burn calories kind of thing. I think I tried to get, because my goal was 60 minutes on the active ring, because you had the step standing on the day active and then calories. So I just tried to get that done. So I think I was trying to go out for an hour. And then one of my friends rang me and said, are you in the flat? And I was like, no, no, I'm not. I'm on a walk. And she was like, I kind of need you to be there because something's happening, happened and I was like, all right, okay. And it wasn't like me or anything. It turns out like my friend who I lived with, her best friend, he died. And she got that information. And I wasn't there because I wanted to go on a walk to get my steps in, which was so unnecessary. I didn't need to do that. If I was there, I could have been there for her. I was there afterwards, after the fact it happened, but she'd already had the information, was a complete mess and didn't have a friend to rely on just because I wanted to go on a random walk to please my Apple Watch. That's how I think about it. And I know there's so many different, like, so many other things could have happened that I could have gone home for the weekend and not been there. Like, and I would have probably beat myself up in the same way. But I think for me, that was the turning point where I was like, right, I need to stop with this because I'm just, I'm being, I'm being stupid now’ (P11).*  *‘It's crazy what it did to my mind as a young person and then into my adult years as well’ (P11).*  *‘Getting a device like this at the age of 13, 14 into adulthood really did take a piece of my childhood away from me because I was so obsessed and it really did create those unhealthy behaviours. And I just, yeah, I just kind of have a bit of hatred towards it sometimes when I look back’ (P11).*  *‘I mean, this is probably a lot to put on an Apple Watch, but I really don't think I would have had an eating disorder if it wasn't because of it. Like, I don't, I just, I wasn't interested in that sort of thing before I got the Apple Watch. Like, I didn't know about it’ (P11).*  *‘You shouldn't give up watches to young people, I don't think at all. I think it really walked my perception of exercise and the reason for exercise and food intake and eating the right food, all of that kind of thing is really important and it's a good, it's good to do it in healthy ways, but doing that to a teenager, I would say a teenager, yeah, teenager, like early, early teens and giving someone that and all of that information. They're only going to take it one way or the other way and obviously I took it the complete wrong way and the awful way, but... Yeah, I don't think, yeah, I think there should be a cap on it’ (P11).*  *‘Girls, especially with body image and as you're getting older, puberty, everything like that, I feel like we have enough to deal with. And then you just kind of chuck in a smartwatch as well to be like, oh, that's how you track it all. And it's, and I don't know, I should say it's a downward spiral, really’ (P11).*  *‘I really am a completely different person. A lot of people said that. To me, everyone thinks it's uni, everyone thinks uni's changed me. And then I've come back this like, great person. I'm like, no, it was the Apple Watch . No one knew how bad it was’ (P11).*  *‘I saw every day as like a bit of a competition. But that meant I was… I was turning down social plans like going to watch a film or, you know, um I don't want to do this because it's going to be 4 hours of driving in total and that's a big chunk of my day. Weird working out logistics like that and deciding if to do something almost based on the window I had left to keep beating my scores, essentially’ (P13).*  *‘I think my wife had been noticing for a while and, yeah, after a while she mentioned it and I was in, I was properly in denial. But she was saying, you know, you're almost, you're choosing to have more time to move over, seeing your kids and seeing some of your best friends and, yeah. I think I was moaning at one point about like going on holiday with two of my best friends because they like to relax and they like the holiday to be downtime and I think that was when she said, you know, I think you've lost your mind a bit. And that broke me a bit because, well, they’re my best friends! I love them to piece and I was genuinely hand on heart not impressed to be spending time with them because I’d get less steps in that day. I fills me with anger now or, or even a bit of shame, because there will be times I said no to seeing people from this, and like I said, some of my friends are no longer with us now. Yeah, I was just a bit blinded by it.’ (P13).*  *‘I think they’re great bits of kits, but I just ended up, prioritising it, prioritising it over other stuff. And it was, it was like, okay, you've had this one wake up on your health, a wake up call on your health, sorry, but what's the next wake up call going to be? Are you going to have missed your last time to see somebody because you were going on a walk, but not even a walk you were really wanted to do. It was wanting to get your steps in’ (P13).*  *‘My wife is often right. She’s also calculated. She weighs things like this up and waits until she’s confident about something to talk, so I hate that she must have been weighing it up for a while, literally knowing I was in this bubble. But I think, I thought at the time, well I don't think it's an issue’ (P13).* |
| Reconnecting with the self | | |
|  | Bodily intuition  (In when to move) | *‘I don’t have a strict set routine. I do this on this day, this on that day. It’s kind of just how I feel’ (P1).*  *‘I had a constant reminder before, and now I’m less distracted by technology. Even though I thought I was looking into my body more, I was actually looking at a screen and now I feel like I’m more relaxed and I can work out because I want to and not because I’m told that I should be’ (P1).*  *‘I was looking at a screen instead of asking myself how I felt before, and now I’m back to being like, oh, I’m a bit tired today so I won’t do that… I feel like it’s made have to listen to my body again, which can only be a good thing, right?’ (P1).*  *‘I probably work out less, but I do feel like it’s goof for my mind because if you were only working out before because you got told that you should but didn’t want to, is that good? I mean, I know physically working out is good for you, but I don’t know. For me, I want to just do it when I feel like it now’ (P1).*  *‘You know what I learned from this? I think I had to learn to trust my body again, which sounds sounds huge, doesn't it? But I got so used to looking at a screen when I took it off I was I was questioning like how how am I going to do this and how will I do this when actually I didn't wear a watch for like two years of my life and managed. But that was that was really strange. So that must mean I was dependent on it, right? That was, that was eye opening’ (P1).*  *‘I do think I probably work out a bit less because I haven’t been given these like hoops to jump through on my watch’ (P1).*  *‘I'm actually quite enjoying it and I'm having to listen to myself more, which I think is a good thing, especially given the context of my weekend run’ (P2).*  *‘Initially when I stopped wearing one, I pretty much, it might also sound quite bad or quite extreme, but I kind of didn't exercise for a good amount of time. I think I tried to pick up running here and there, but I couldn't ever get into the actual routine of doing something, but I think then I was probably using the watch as a bit of like a crutch as like my actual reason to exercise, whereas I wasn't wanting to do it like for myself’ (P3).*  *‘At first it changed my behaviour because I stopped like exercising… almost like in a way that I just don't think I like wanted to or like in the headspace I was in and the kind of part of life I was in, I didn't, I didn't again feel pressured to do it anymore. So I was like, I'm just going to do what I want and then I think I've now come around to, and like I tend to be on my feet and quite active at work anyway’ (P3).*  *‘I now like have started kind of in the past few months exercising again, but because, because I want to, and because it's like good for your longevity and body and things and brain more than anything really, but not because I feel like I should’ (P3).*  *‘It's just like I'm living a different lifestyle now. I still like hiking, I still like swimming. I've slowly picked up all of the activities, but it's can you fit this into your busy life instead of I need to do this on this day to like get this score’ (P4).*  *‘I'm not just seeing it as this what's the word, maybe like test subject or like product’ (P4).*  *‘I think I probably am like slightly less active just because I'm not being like prompted to do anything. But I think part of the reason that I bought this because, you know, like it was in lockdown, I was trying to get activity in, I was trying to lose weight, all of this. So it's always been kind of prompted by like concerns about, you know, like my weight and my appearance. And I think I've also been trying to make like a more intentional shift to like doing activities that I enjoy when I want to, and I think that the watch doesn't really fit into that vision’ (P5).*  *‘At the moment I'm trying to just be a bit more mindful about like my approach and doing things when I like where I enjoy them and stuff and yeah I just don't think that the watch helps me with that’ (P5).*  *‘It's more about satisfying my needs rather than like hitting, like you said, like the thumbs up or the big run X. It's more about like, what do I actually need and like what is actually helpful to me in this time’ (P7).*  *‘I remember like pushing through illness and things because, yeah, I, I thought it made sense’ (P8).*  *‘My wife asked me about it because I said that I was going to do this. She joked and was like, it's like you're not checking in with your boss like every day. It was like it was like a micromanager . And it was, yeah, so a relaxed is what comes to mind in that specific area. I'm still busy, I'm so stressed about other things, but it definitely removed a bit of pressure there’ (P8).*  *‘Before I was almost thinking of it as like this, this employee that had this to-do list every day and like had to do it. So I am more chill now in terms of health … I feel like I'm probably just a bit more relaxed. It's not that I'm being lazy, but I'm weighing up like, should I do this workout today or tomorrow? And I'll be like, oh, I'm quite tired today. Whereas I would have looked at data before and said I could do with doing that now or okay you don't have to do it today. So it's like I'm making the call now instead of my ring or watch. I think I'm probably just making the decisions a bit more. Yeah, instead of listening to what I should do’ (P8).*  *‘Before I might have been really tired and I would have looked at my data. I said no you need to do it and now instead I'll just I don't have the data so I'll say I'm actually really tired today, I'm not doing that. So I don't think it's changed like levels but I think it's changed like how I approach it’ (P8).*  *‘I'm thinking about AI right now, this concept of, you know, we're going to have a generation of people that won't think about things in depth because we can adopt the mindset of why I would do that when a quick search on ChatGPT is quicker, like it's quicker, it's easier, it's efficient? It's trying to be smart with time and actually you are not doing the thinking yourself and I think, without realising, I just, I wouldn't ask myself, I'd just look at the watch because it could tell me… Instead of how has my day been? How do I feel about that? You know, asking myself that’ (P8).*  *‘My mindset without realising was in a bit of a confused point of view, saying I'll just, I'll just look at the app instead. Let's just see what my data says’ (P8).*  *‘Since then, now I'm in control. I'm not like, I'm not being a slave to the watch. The watch has been a slave to me’ (P9).*  *‘I look inside and see how I feel. I ask myself how I feel’ (P9).*  *‘It's all the health, the health thing, or it says, oh, you need to rest for 72 hours. I'm like, no, I'm going to go for a run tomorrow. So it's, I mean, I'll ignore you’ (P10).*  *‘I definitely wouldn't choose to wear it if I knew I was having a day where I was sitting down a lot because that would then make me think about it and I wouldn't want to think about it because I feel like it's still ingrained in me now, every hour’ (P11).*  *‘I'll go and exercise when I want to go and exercise. And I'm doing it for me and my own health and fitness. Not because a watch is telling me that I've only burnt so many calories today so I need to go burn some more’ (P11).*  *‘I would say motivation wise, I'm definitely not as, I'm not as motivated to exercise. I will just, if I want to exercise, I will, but I'm not going to. I don't find myself waking up every day and thinking I'm going to go to the gym… whereas it was when I was with the Apple Watch, it was, I'm going, I'm going to, I have to go’ (P11).*  *‘I just felt that, yeah, I was paying too much attention to an app rather than my own body. And so it just didn't feel very like genuine. Like I was exercising because my watch told me that I'd had enough recovery and I should move. And then, yeah, and I was like, this doesn't really take kind of, it doesn't ask you how you feel. It doesn't ask you for your kind of feedback, it just sort of, and so yeah, I just preferred to go kind of based on how I was feeling on the day’ (P12).*  *‘I think I'm probably a bit less snappy with people. I'd look at numbers before and then I'd be working out how to fit this in and how to fit this in. I don't think I'd explained that to others, so I found myself just getting quite snappy when plans actually were flexible, I just I just made-up these rules, and I think that came down to just feeling, feeling like I had to meet these goals… I just saw it as this thing on my to-do list that had to be ticked off, non-negotiable’ (P13).*  *‘It's having that quieter brain and it feels better for my mind. I've got less of this urgent to-do list? I've still got a to-do list; I've still got urgent things on it, but I think I was putting movement at the top of it before. Now it's still a priority, I'm still very active, but there's almost there's less of a telling off if I don't do it, that's how I'll put it. There's less of a, like, yeah, less of a telling off I give myself if I don't do it’ (P13).*  *‘I'd say it's not actually changed, not changed what I do on paper because I'd already formed that routine. I'd used the watch to help me form that routine and it was absolutely brilliant for that. It's more like the mental processes behind it and you know, why am I doing it? Do I want to do it or is it just on my schedule today? If I don't want to do it, can I swap it round?’ (P13).*  *‘So it's given me more independence in that sense…What I do, but independence in my thoughts really, to then decide what I do and when I do it. Before it was like I was making decisions for a collective or me and the watch, which sounds stupid, but numbers were always in mind’ (P14).*  *‘I weigh things up a little bit more… just thinking for yourself instead of thinking for you on the watch, because it does become this combined combined mindset, really. It's not you versus you watch. It's like you're a team’ (P14).*  *‘I think I'm doing less like formal exercise, I've still got my one class I always go to, but some days I would do like longer ones because I'm looking at numbers and I think I might be doing less but it's not necessarily a bad thing. I'm doing it. I'm doing it because I want to now, whereas before it was a bit, I don't know, I just, I would prioritise it too much and then be sick of it a week later’ (P15).* |
|  | Greater self-acceptance / less critical of self | *‘I’m aware that having that data, I just started being so critical. I needed reminding I’m just a normal human being, like trying to get 1,000 steps in. It’s not that deep’ (P1).*  *‘I found myself really beating myself up when I got busier with a job and and when I got ill and I'm still ongoing trying to get like a diagnosis, but it to me I feel like I have, I like have flare ups of fatigue I think so I already had to do a lot of like forgiving and mindfulness around that, a watch telling me to move when I couldn't was the last thing I needed. It genuinely made me angry and it got to the point of me questioning. Hang on a minute, what, like, why is this on my wrist still?’ (P1).*  *‘Having that data to be more competitive means you you can zoom in a bit too much and focus and think, oh, I just missed this. How did I not do that? Oh my, I felt a lot quicker on this stretch, but actually it was this. Whereas because I don't have that even though some days I'm really bitter that that is gone because I do enjoy looking at it, it makes me just appreciate what I've done without getting into the nitty gritty’ (P2).*  *‘I think the time I had off from it, I think I was also just in like a space online and things like that, which was kind of being about not like having to exercise. You don't have to force yourself to do it for a specific body type, a body positivity kind of like lens that I think I grew into a lot more during that time where I wasn't wearing it’ (P3).*  *‘If I didn't have an active day, I’d feel rubbish’ (P3).*  *‘Another time after that he said, seriously, I don't think I've seen you look at that watch and look happy from whatever it's telling you’ (P4).*  *‘It's interesting. I can't really work out if... because I think me trying to like be a bit easier on myself kind of led to not wearing the watch as much. So I can't tell how much not wearing the watch has like enforced that, yeah. I do think it probably has helped. I think, yeah, there were just some times where I would just feel so, just so irrationally like down and hard on myself because the number on this like stupid watch wasn't right’ (P5).*  *‘I'm definitely less active but I don't know the extent to which wearing a watch has made me less active or, you know, in part the reason I took off the watch was because my like circumstances have changed and it's stressing me out to like hold myself to standards that no longer work’ (P5).*  *‘I definitely feel less stressed. I feel like when I think about like my activity, my health, my like body image, I just feel like I have a more pragmatic view now’ (P5).*  *‘If I think about my like watch usage, it's always been connected to like my own like body image and things like that and like activity as a way of like managing that, I suppose. And I think, yeah, so I think that like process of taking it off is kind of, for me, part of like a more intentional. It's control, really, just like trying to change how I think about these things, which is obviously like very, very difficult given society’ (P5).*  *‘There wasn't a moment where I was like, right, I'm taking this off. It was more just like a gradual, like trying to be more accepting of the fact that like, you know, A, my life has changed, like things that were reasonable, no longer reasonable, but that doesn't have like some, you know, like, good or bad, like moral bearing on like my body or me or whatever’ (P5).*  *‘I do struggle a little bit like with like my weight and things like that. And so I think it was just a way for me to like kind of bully myself further by like, oh, you are this way because… I think that was just a way for me to like justify my negative emotions towards myself. Like it was a way to like put a numeric number on it, like, oh, you are this way because you didn't hit this metric. And so I think removing it has sort of like, I'm very neutral about those things now. I don't really fixate on them as much’ (P7).*  *‘The first thing that shot into my head there was like body image and stuff. I'm not suddenly like a different person, but I do feel, I think I'm thinking about my body less’ (P8).*  *‘With MS, we get these relapses, you know, so we get moments in which, well, I do get fatigue, where, you know, it's not very nice. It's not really helpful because it does nothing but remind me of the condition that I'm in, you know. So it doesn't really help’ (P9).*  *‘If I started tracking, say, my walk to the office, then that's unnecessary. I don't need to do that because who is that for? Because I kind of, I feel like I sometimes hold it as who am I doing this for and I need to remember that I'm doing it for me now. There's no person who's watching me. There's no one judging me for it’ (P11).*  *‘It's actually changed more positively because it's I just feel a little bit more relaxed and so there's just a little bit less like negative self-talk. If I've maybe not hit a goal or if I've done an exercise and being like, oh, that only burned so many calories. I think not knowing that, yeah, not knowing that is like, okay, well, like, that's not really what I think exercise should be about. I don't think it should be like a sort of means to a reward or a means to sort of punish yourself. Like it should just be a nice thing to do for your body’ (P12).*  *‘I was seeing it a bit less of a list of things that need to be done every single day and I have zoomed out a little bit. I'm doing my strength training. I'm getting a weekly average of more than enough steps in, but some days might be quieter and some days might be more active, and that's the ebb and flow of the working week. That's the ebb and flow of, you know, being tired this day and needing to recover. That’s normal. Tired isn’t bad, but I saw it as something to deny and ignore when I was wearing my watch. I saw it as a, a constant thing to tick off and saying I need a day off before would stop me ticking things off. So yeah, I think, I think it's made me a little bit more... understanding that I need rest and that rest is as productive as movement. It should be a balance’ (P13).*  *‘I just feel a bit, a bit nicer to myself. I think the reason I took it off was because I was so focused on these numbers, even though... No day on in trails is the same. You could do the same route and it could be 3 degrees hotter. And it could be a complete different day, or it could be the same temperature. But the wind direction has changed and it can be a complete different day and I was ignoring that and essentially... I was bullying myself. It was me doing, doing the bullying about saying this isn't good enough. But it was the, it was my watch whispering the things in my ear to say out loud, you know, if I was in a gang. They were fuelling it. But I was someone saying it to myself’ (P14).*  *‘I think this is my favourite training block I've done. It just feels a bit more gentle. You've put in all the work, however, it's just been relaxed. It's felt, it's felt kinder and calmer, which is, I suppose, exactly how it how it should be given. It's just a passion project, friendlier. Yeah, just for joy instead of any punishment from your wrist’ (P14).*  *‘I feel a bit less, like, frustrated by myself, I would beat myself up some days for essentially being bang average, when average is normal. That's the whole point of average’ (P15).* |
|  | A sense of freedom | *‘I just feel quite, it’s weird to say peaceful, but I think that’s the best word I can think of to describe it’ (P1).*  *‘One of the reasons I took it off was because I just got a bit into this idea of the fact we need to improve at all times and I was definitely on the road to burn out from this watch, so I kind of don’t monitor things now or try and not to, and just chill which not having a watch allows me to do’ (P1).*  *‘They just made me a bit too obsessed with getting better all the time so I much prefer not having more and I think it it gives me like it makes me slower but I mean not in the fact that. I've started trying to have like a slower life at a slower pace and and like more relaxing and I think taking it off is something that's really helped me’ (P1).*  *‘The first thing that came to mind was, like pretty free, it's very freeing’ (P2).*  *‘I am enjoying it. I said to my partner, it's like I'm just like, I'm just like a mountain goat and I'm just trotting around’ (P2).*  *‘It's making one run a week completely peaceful … because I don't know if you can fully switch off when you're looking at your watch that regularly’ (P2).*  *‘It probably made me think about [my body] less and like actually just not view it as something which I had to think about in quite the same way, because when you're constantly kind of like faced with all the numbers of how many steps you've walked, how many calories you might have burned, how many minutes you've been after that day, I think you think a lot more about your body and what effect that might have. You become a bit of an object, and then I think when you remove that, it was just like I didn't have to think about it all’ (P3).*  *‘You get used to it and you're like, wait, but do I actually enjoy it? Is it good like for me? And then you get to an age where you start to question it and you're like, oh, hold on a minute. Maybe it's not’ (P3).*  *‘It went from these nice messages of what I perceived as encouragement to like heckling… it was telling me that everything was down as well. The sleep scores I got in the morning were bad and I thought, hang on a minute, it's only going to get worse. It became negative and... It was this weird, like, I do care, but not right now’ (P4).*  *‘It's not telling you anything you don't know . I know I'm tired, I know I'm worried, I know I'm stressed, I know I'm not working out as much as I was before. The only thing it's bringing me is like feeling bad about it, so I took it off’ (P4).*  *‘The watch after baby and during baby made me just feel bad about what I was doing so taking it off felt like a bit of a relief. It felt like I'd put my phone on silent mode and I'd like I blocked it out, I put like noise cancelling headphones in. My behaviour hadn't changed but I wasn't getting told off about it. So it felt really nice. It felt, yeah, it just felt like the logical move. I don't think it actually made me happier, it just removed the frustration I was having about my watch’ (P4).*  *‘It was just the daily, the daily notifications to move and stuff. I think what I'm trying to say is I love my watch in the moment for wearing it for exercise. I don't like my watch the rest of the time when I felt like it was nagging me and not recognising like I've got a child. I need to do these things’ (P4).*  *‘It's reminded me that like, I'm just a person who's got responsibilities and... Who likes exercise and who is who is actually really fit, but that's not the only thing I have to do, you know, you get a notification to move, but you don't get a notification to meal prep for the week. You don't get a notification to do the washing, to cheque on my sewing that like you don't get anything else. The only thing you're really getting told to do is to be fit and I think that you can you can have blinkers on it, so it actually may give technically had a negative change in how I view my health because I see it as, yeah, I technically see it as less important than I previously did, but that's because I think I've got a bit of a better focus on the rest of my life as well’ (P4).*  *‘I definitely do feel just like less stressed about my health and like less neurotic about like getting the boxes and like ticking the boxes’ (P5).*  *‘I don't worry as much about like, oh, how, like, what's my heart rate going to be just now? Or am I expending enough energy like doing this exercise? I just don't think about it whatsoever. And it's really nice to not be like, second guess or try to predict like, oh, the watch is going to say X amount of calories or heart rate’ (P6).*  *‘It slowly consumed my life’ (P7).*  *‘No matter how arbitrary they were, I was concerned with my sleep score. I was concerned with my like, and I was spending more time really concerned with the aspects of like hitting my step goal, hitting my caloric intake, hitting my sleep goal, hitting all those metrics, no matter how good they were for me, I was more concerned with the number than the actual like benefit that they had for my life. So I was just like, yeah, it was it was like my my perfectionist genes. I just wanted to do really well at something’ (P7).*  *‘As somebody with ADHD, I find things to fixate on. So like, it's not, removing it was like a good thing because I'm not fixated on that’ (P7).*  *‘My mental health overall has like kind of improved because I'm less like, I don't feel as bad about not hitting those metrics because I don't know what the metrics are . But I think like overall, not much like physically has changed, like in terms of like activity level or anything like that. Yeah, I would say my mental health has increased since removing it, because I'm not as like fixated on those numbers’ (P7).*  *‘I think that the metrics that it measures, I think that the metrics that it measures like still are important. I think that it is like, like I still value those aspects and I still think that they're very important to an overall healthy wellbeing. But I think at the same time, I think it really did change, like…how much pressure I was putting on myself to hit those metrics’ (P7).*  *‘It removed a lot of the kind of anxiety I felt about like not getting better all the time and like not doing more each day’ (P8).*  *‘I just became aware that I was avoiding getting the data because I knew I hadn't done well. And it wasn't like it was really stressing me out. I was putting pressure on myself, to, you know, like go for a walk, go to the gym’ (P8).*  *‘I'd tell myself to just put it back on, like I'd say, it is actually quite good for your health, put it back on. But I just felt like so relaxed. I just didn't think about putting it back on’ (P8).*  *‘If I wasn't moving, like every time, it would send you a buzz to remind you, move now, you know, and then it created anxiety in me because I'm like, I can't move. I'm doing stuff, right? So I took the notifications off as an attempt to regulate’ (P9).*  *‘That's an additional thing for me to worry about. I'm like, I have enough things to worry about. I don't need my watch to tell me that I'm not sleeping well. I know’ (P10).*  *‘I realised there was much bigger things going on and I am obsessed about a little Apple Watch and what food I'm eating every single day. So that's when I kind of, I don't know how to describe it. It feels a bit like a jail, like I was in an Apple Watch jail’ (P11).*  *‘I feel judged for my own Apple Watch. That's crazy. So if I do, if I don't know, I don't know how to describe it and I don't know if I'm just really strange, but say that I was about to go spinning and I was like, oh, I'll get my Apple Watch. And then like, I get that like pit in [my] stomach’ (P11).*  *‘I remember that week, I didn't go to the gym. I didn't wear my Apple Watch. I just lived my life as normal. And then that's when I was kind of like, oh my God, living in this tunnel of Apple Watch-ness and like trying to please someone who's not even a person, for what?’ (P11).*  *‘I feel so much better. I'm so much happier, so much healthier. I just, I actually, like, I don't, this sounds really, really stupid, but I really don't know where I'd be if I was still in that state of complete and utter obsession over something. Yeah, I don't, yeah, I don't know. Just, just free. Like I've, like I said before, it's kind of like, it was a jail and I kind of stepped out of the jail and then I like, say that was the bars and then I went into like, just like the police station and then I went outside, was at the point where I completely stopped wearing it’ (P11).*  *‘The more I wasn't wearing it, the better I got with it, with I don't need to be doing this every single day. And then that's kind of when I started to slide into my kind of routine and it got a lot easier. And then now I don't really think about it’ (P11).*  *‘I was just trapped in it, I would say. And the only way I can compare it is, you know, when you hear about how if people have a problem with alcohol and they go to maybe different petrol stations every day, so people don't know that they're drinking as much and they hide things under their bed, that kind of thing. I feel like that was me with my Apple Watch, like I do other workouts between my workouts and not show people those workouts, but look, I just burned a lot of calories in the one workout. Does that make sense? Like I was doing things like that and I was like, that's why I kind of related to an addiction’ (P11).*  *‘It definitely felt more kind of freeing and I was able to enjoy the activities more without kind of treating them as an end to, like a means to an end or like a way to like meet a certain target sort of thing. And so I think it did change the way I view exercise a little bit. And I don't think that it affected how much I exercise. I think I was able to be more consistent, but kind of less dreading doing stuff’ (P12).*  *‘I just feel a little bit more relaxed towards exercise and just towards kind of getting up and getting on with my day. I feel like it's one less thing to check and sort of less data to be kind of going around in my brain’ (P12).*  *‘It was just that changing pressure and like guilt and watching the time and thinking you've only got 2 hours left to do this that changed’ (P13).*  *‘It was a constant back and forth on numbers in my brain, so I’m just a bit more relaxed’ (P14).*  *‘I see my body in a very logical like machine way for fuelling for runs. Calories in, calories out, need this many carbs works well for me. Like you get into a routine and it becomes very structured and objective. But I think I was applying that same logic to moving and to my running, but also to like everything was, oh, can I get a little bit more in there? You know, I think some days I should have just driven to a shop and I was walking just because it's a bonus of steps, which is really good, but it comes to it comes to a point of, come on now, you're wasting time. And so I think taking it off has made me less, feel less pressure to always be thinking about the numbers, the numbers, the numbers. Which has been really nice’ (P14).*  *‘It does become this combined mindset, really. It's not you versus you watch. It's like you're a team. Yeah, you're on the same team and wanting to get better data. So removing that has meant it's just me. So I'm seeing myself a bit more as a just normal person instead of a machine’ (P14).*  *‘I'm a bit more relaxed in what I do day-to-day’ (P14).*  *‘I do think my behaviour's stayed pretty much the same, it's just someone's just not shouting in my ear the whole time’ (P14).*  *‘I think this is my favourite training block I've done. It just feels a bit more gentle. You've put in all the work, however, it's just been relaxed. It's felt, it's felt kinder and calmer, which is, I suppose, exactly how it how it should be given. It's just a passion project, friendlier. Yeah, just for joy instead of any punishment from your wrist’ (P14).*  *‘I can't decide if it's helped or not because I think I do think I'm probably less active, but at the moment I seem relaxed, it does seem to have fixed me getting a bit stressed and down and I have been, I mean, I've been less all or nothing. I've been reduced, but I suppose it's that like sustainable working out when I want’ (P15).*  *‘I'm a little bit torn at the moment with this whole like, I feel more chilled out, more relaxed, but I'm also not working out as much’ (P15).*  *‘It was pressure to be fair. It was this weird, like, nothing bad's gonna happen if you don't do it, however, your numbers are low. It’s kind of just like awareness. A bit like, um if you're doing your job and you're aware that they're monitoring you at the moment. It was just, it wasn't this huge pressure to completely change, but if you were debating one or the other, you'd go for the better option with the watch on. Just aware of some kind of responsibility. Which sometimes I loved but other times made me feel a bit… itchy, a bit like I had to drop everything to stand up’ (P15).*  *‘The reason you buy a watch is to be reminded about your health and put your health first if you've not been doing before but I would argue that in some cases, it doesn't need to be first, because I will tend to put it first and then neglect everything else and then put it eighth. So I'm hoping instead of it being first, it's just I'm aware of it. It's a priority, but it's not top priority. I think it's just making me a bit more balanced which was my goal’ (P15).*  *‘The pressure from it for five years, I think it’s that that wore me down. Maybe, like I don’t know how it’d ever be a thing, but short-term use, like renting it for a month to just, like, learn enough about yourself and then you’ve got the information without this weird feeling’ (P15).* |
| Difficulties in habit reversal | | |
|  | Removing external praise brings disappointment | *‘I loved it. It was this cheerleader and it’d be like yeah! You’re doing amazing! Like, keep going! It was really good for me and I think now I’ve stopped using it I don’t want to admit how much I did really enjoy that, but I did’ (P1).*  *‘Doing a really long hike, I’d be annoyed I couldn’t see how many steps I’d done and like how long I’d been out there for and stuff. At the start I was so disappointed because I didn’t have the watch telling me I’d done good, but it’s pretty normal to me now’ (P1).*  *‘Now I feel like I probably get less enjoyment because I'm not getting that praise. I'm not getting that little like well done, you did this, but for every like bit of enjoyment and feel good there was that there was like ten more bits of competitiveness and stress, so I think I'll survive without the enjoyment’ (P1).*  *‘There's glimmers of time when I miss it when I know that I've done good and could have had it like recorded’ (P1).*  *‘Even though it sounds doesn't sound a lot, it doesn't sound too impressive. For me, not having my biggest run recorded, it almost felt like I was fobbing myself off or something, so yeah, it was genuinely was big for me’ (P2).*  *‘I actually felt like really, really fit and fast there and I would love to know what I did on this section and this section because I do love looking at the breakdown at the end like it, I just really enjoy it. And if you're running for like two or three hours, that's a big breakdown you can get into. So I sometimes feel a little bit like, I don't know what the right word is, maybe a little bit frustrated, like just a bit, a bit grouchy for a second thinking why didn't you do that? But I've also tried tracking it but not looking at my watch and I just I just couldn't really do it if it's there and on I, I do look at it so, yeah, I still think taking off is the best option, but I get back after a run and think oh, I actually would like a bit more of a detailed breakdown there. And I did like sharing like what I've done. I used Strava a lot and I can't help but think that it's not being counted in my totals’ (P2).*  *‘It's not there to share and I actually really enjoy sharing what I've done on Strava and I always take some pictures and like write a little a little post about it so there's that element as well. I don't think it takes away from what you've done, but it's almost a nice little reflection at the end. If you have the data and the pictures, it's like a little a little summary of what you've done (P2).*  *‘I'm really competitive and I think that's why I stick to my plan and I do think being competitive strengthens your running. It means you push yourself more’ (P2).*  *‘Strava was funny because you're showing people what you've done. I don't want to say it's boasting, but it is drawing attention to you. It's saying, look at what I did this Sunday morning, and so it was a bit of a second layer of reward, like people giving you kudos, they're being like, yeah, fair play like this looks ace smashed it. You are removing some reward’ (P2).*  *‘Sometimes it is nice to know numbers and to know if you've walked a lot that you have done however many steps is a lot for you’ (P3).*  *‘You look at the watch and sometimes like, I feel like there must be like a dopamine rush type thing associated with being like… yeah, you almost don't like, unless you're going with like a friend who's saying like, oh, well done, like you managed to lift that or whatever. You don't almost have the same immediate feedback and like a kind of, I can't think of the word, but yeah, you don't have that like confirmation that you're doing well’ (P3).*  *‘I think I missed the data about like sleep and your sleep quality. I could be like quite interested in like knowing more about that because apart from kind of when you wake up in the morning, I feel like I never have an understanding of how I've slept, whereas that was really interesting. And I remember that was kind of one of my favourite parts of the data’ (P3).*  *‘When I did get to work out every so often, I thought it would it would be nice to know what I'd done’ (P4).*  *‘I used the watch as like a record. It was just a record of what I'd done that you could look back on. So it was weird and I would say it was dramatically negative emotions, but every so often it was a bit of a like, aw, I would have liked to know that’ (P4).*  *‘This is the bit that I kind of really do miss. Sometimes I don't care, you know, about activities, but others, specifically swimming, I can't count the laps I've done in swimming. I zone out a lot in swimming. And in running, I might know estimates of routes, but I don't know things like speed. I'm not, I'm not angry about it and things like that, but there's curiosity there on what I've done’ (P4).*  *‘I feel like sometimes I use my watch to force me to push harder, like it was. I was already doing the workout but it was a bit of motivation to push harder because like my heart rate was not at a certain level and things like that. So motivation, you're relying only on your brain then. You don't have something prodding you’ (P4).*  *‘One thing I don't like is that I miss up on the like annual summary, I paid for Strava, I used it a lot and I really loved it and I feel now I don't have the data for it. So actually if there was some kind of way for me to have Strava because it's like my little diary of what I'm doing’ (P4).*  *‘It would tell you like, hey, it's been a while since you've moved. So I don't get those indicators. And because I have ADHD, I will get sucked into an activity and then not move for hours at a time. I think that was the one thing I truly do miss about the Oura Ring is those indicators that like, hey, it's been a minute since you've moved, maybe you should get up and do something’ (P7).*  *‘I do like going on big hikes an looking at that data and like, oh, what did I do? Because I know what I did, but you know, if you've been out for seven hours, you zone in and out. I can't tell you the steps I did and the time it was moving and my pace. I just can't tell you that so l might be sad in the summer about that’ (P8).*  *‘I'm enjoying it and I might be less motivated in the moment to like push more but I think that that was from removing the screen’ (P8).*  *‘If I went running and I didn't have it, I would feel a bit disappointed that I can't check. But then it happened, it happened several times that I went running. I'm like, oh shit, I forgot my phone. Can you imagine if I go back and get it? Not a chance, I'll go running. And if it, you know, and I'm like, well, the run still happened. I still had fun and the stupid watch doesn't even know it’ (P9).*  *‘If you get so used to tracking something and I was tracking it on my watch, when you stop doing that, it feels like you've got no evidence of what you've done. It's really hard to explain it to someone that hasn't used a device like my children have, so they completely understood what I said. But my partner, someone used to watch, doesn't understand it. And so we had these huge debates, and I want to say it was a debate, but it was two people stating their views. There is no understanding from the other person. When I was saying, your recordings become proof that you've done something, it becomes like a little logbook. And so, when you have all of those, and then you should only stop, it does feel like you're not gaining the same thing from it. I don’t know. Yeah, like this was proof of what I've done before and it was like you got a little certificate or a medal at the end of doing something and then all of a sudden you don't have that. And they did, it felt really different and I think it's what I hated the most about removing it’ (P13).*  *‘I'll see people tracking things and every so often without sounding stupid, I do feel a little bit envious. I do look and I think, oh, I won't want your having, that's nice. But family comes first’ (P13).* |
|  | Recognising behaviour was compulsive | *‘The data was there so I couldn’t help but look at it, which sounds like no self-control, but actually when it’s all in front of you to like such detail, it’s hard not to look at it. So I looked at everything, I really did’ (P1).*  *‘I kept going to check my wrist and thinking what have I done? How am I doing?’ (P1).*  *‘I still go to check my watch at the end of a run. I know I'll still go to, like, stop tracking on some weeks’ (P2).*  *‘I tend to just find myself looking at my screen. I've not intended to and I'm on my watch’ (P2).*  *‘They don't really realise that you don't have to and you don't need it. I think we're honestly quite too used to having technology around us’ (P3).*  *‘My mum who wears one 24/7 has a lot of like problems with body image and feels a lot of kind of negative emotions if she doesn't exercise and doesn't hit her goals and she sat on the sofa like pretending to her watch that like she stood up and things, and I think I used to remember like, yeah, walking about the living room in the evening, trying to hit the step goal’ (P3).*  *‘Using my watch was just like the normal thing to do and it was automatic’ (P4).*  *‘I think that it kind of just reinstated my hatred of technology, I guess, especially with like the advancements that we're seeing nowadays with like AI and stuff, like how it's integrated into every aspect of my life’ (P7).*  *[Discussing moving from smartwatch to ring] ‘Yeah, like you still get notifications to your phone, but it's there's a different level of like investment almost. You're not checking it the same as a screen. You see how you’re doing out of curiosity and then you don't have a screen again, essentially. So I enjoyed that. I actually think that might have been without realising wind down in wearing a device’ (P8).*  *‘You build up a streak and obviously it's the higher the number, the more you don't want to get rid of it, I can't remember what I ever got to, but I know it, I know it was stupid’ (P8)*  *‘Before I took it off, I felt a bit like, silly, I thought oh, you can, you can just ignore it, but if it's there on an app, I'm not ignoring it. And so I felt a bit it feels like this is an intervention, I'm taking it off you. You know, like someone saying, just don't eat chocolate in your fridge instead of like don't buy it at all like it felt like a weird self-control issue with me, and I did, I did feel a bit silly’ (P8).*  *‘I find myself doing that and I'm like, oh, I don't have the watch’(P9).*  *‘I didn't tell them that I was obsessed with it because at the time I didn't even realise I was obsessed with it’ (P11).*  *‘I feel like it's still ingrained in me now, every hour. I get up and I'll walk somewhere. And I've noticed it since starting my PhD because it was always, I never sat down for like a long period of time, never had an office job before. And I do honestly, it's every hour. It's kind of on the clock really when I first do it. So say I came in at half eight, 20 past nine, I'll get up and go for a walk and then 20 past 10, then 20 past 11. Like, it's quite routine and it's, I don't know, I don't know why. Maybe it's from the smartwatch in the past of like knowing every hour it tells you to stand up’ (P11).*  *‘I do think if you're a person who reads into things and likes data and stuff like that, that's when it gets just really obsessive, I just call it. Like it's, and I've not really gotten, I don't think I've got a really obsessive personality, but there's something about a smartwatch that really makes me, like, I'm just, I'm constantly looking at it and you're just constantly, like, you're checking your heart rate, you're checking how many calories you've burnt, you're checking how much activity you've done every like 2 minutes’ (P11).*  *‘At first it was definitely quite difficult. I kind of, I went through a bit of a brief moment where I didn't feel a lot of purpose in my day-to-day because I wasn't, I didn't have anything to show for it, which was a bit strange’ (P11).*  *‘I was just trapped in it, I would say. And the only way I can compare it is, you know, when you hear about how if people have a problem with alcohol and they go to maybe different petrol stations every day, so people don't know that they're drinking as much and they hide things under their bed, that kind of thing. I feel like that was me with my Apple Watch, like I do other workouts between my workouts and not show people those workouts, but look, I just burned a lot of calories in the one workout. Does that make sense? Like I was doing things like that and I was like, that's why I kind of related to an addiction’ (P11).*  *‘I definitely wouldn't choose to wear it if I knew I was having a day where I was sitting down a lot because that would then make me think about it and I wouldn't want to think about it because I feel like it's still ingrained in me now, every hour’ (P11).*  *‘My relationship with my smartwatch. You know, do you know, like when you've like an ex-boyfriend, when you first break up with him, you think about him all the time. And then now he's just like, sometimes I'll think about him, sometimes I won't. But that's what it's like. It's like a, it's an ex, but it's like an ex of like two years. Like I'll occasionally mention him in passing, but never, I'm never thinking about them like constantly anymore. Like that's kind of where I'm at with it’ (P11).*  *‘It's an all girls run club. There's quite a few regulars. So like I've developed quite a good like social group from it. And kind of when I mentioned that I wasn't going to be wearing a Fitbit anymore, they were sort of like, oh my God, I would die without my Garmin or my Apple Watch’ (P12).*  *‘If it was in the house, it'd still tempt me. Like, I felt like knowing it was just there, it was almost like whispering for me to put it back on. So I actually felt that [giving it to their daughter] was the better option’ (P13).*  *‘I had this full, like, adjustment period in crisis because I was so used to looking at my watch. It was the equivalent of like a teenager now, not having the internet and not knowing what to do. Without realising that I'd become me, I was so connected with it all the time. I was checking my wrist so often and every time going, oh God, it's not there, how am I meant to know this? So it was weird. It was genuinely a 180 when I took it off. I must have been looking at it so often cos I just remember feeling lost for a while’ (P13).*  *‘If you didn't do this 3000 steps today, you could physically do it tomorrow, you could catch up but it was having this little, little like sticker on a calendar chart that, it just meant to me that if I didn't do one, it was this glaring, it was almost a big circling, so you know, missed out here. And they work on streaks on Apple, so the longer you did it, the more obliged you were to carrying on. And it turns out hat got to me a little bit’ (P13).*  *‘It was very quiet when I first removed it. I remember thinking like, well, what have I done? What should I do next? So I suppose you might see that quietness as normal and good, but I wasn't used to it, I was freaking out of it with the radio silence because you get used to having this back and forth conversation. It's whether you like it or not, it's a conversation. The watch tells me something I think about it. It tells me something else I think about it. And it was this full dialogue I had on how to complete goals, essentially’ (P13).*  *‘I would do something and instantly turn my wrist and almost have this slow motion realisation again and again and again that I wasn't wearing my watch anymore’ (P13).*  *‘If you asked me a month in, I would have told you I hate it and it's stupid. And that was just this, I was in the middle of this really slow process of getting used to, like changing the way you view something and changing the way you judge something. So at that point I was itchy, I was irritated, I hadn't really felt the benefits yet, I was just missing my watch’ (P13).*  *‘I know that I will be looking at things like time elapse, heart rate, all of that, just because it's there and I don't have restraint, really. If it's there, I'll look at it’ (P14).*  *‘It's interesting, I feel like it's solidified why I did it, and that it was a good move for me, but it's also reminding me of like the good times I had with a watch. It's a bit of a romantic montage because it's this constant connection. So thinking about me getting the data for my race is truly in my head now. It's not like teasing me, I will not put it on before but you get so used to it that, it's oddly (P14).* |
|  | Substitute tracking | *‘I used Strava a lot and I can't help but think that it's not being counted in my totals. But sadly to get around that. I have a little list of what I do on my long runs that I'm physically writing down in a notepad, so I do know what I've done’ (P2).*  *‘I will like track my steps on my phone now instead of like on a watch because my phone is often in my pocket or nearby to me, like on my personal and walking somewhere. So I can still like track things, but it just takes a bit more effort’ (P3).*  *‘A couple of times I've been with my friends and I have asked like essentially, what we've done from from their watch, just to, just to get a bit of that data back’ (P4).*  *‘I can ask someone else what we did if we've been walking together, like if I really want to know’ (P8).*  *‘I downloaded Strava, so I could get kind of that more in-depth detail in terms of like pace and like distance, so I could plan a route and stuff like that, stuff that I could do. And then I realised that I was relying on Strava more and I wasn't really interested in kind of what the Fitbit was telling me’ (P12).*  *‘I still put in like a yoga class onto Strava and it'll just be one hour yoga or one hour Pilates, but it's got nothing to do with like the amount of calories I burned or anything like that. Like it's just not that important to me anymore. And so now it's literally just to keep track of like, say, an average of four things a week’ (P12).*  *‘I was looking at my watch so much, but I had a very, I had a very easy way of judging a lot of my progression, because with weightlifting, it's numerical. It's numbers of reps, it's weights being used. You do have this way to monitor what you're doing. And I mean, I used to input that data to my watch. So now what I do is I just I just make a note. I've got a tiny notebook that I shove in my gym bag and I just make a note of where I left off at to see do I need to go up away or should I stick on this? More for memory over a record of things now’ (P13).*  *‘I've taken it off for all of training which feels hard and a work around because a lot of my training is saying be out for this time with this elevation. So if I plan a route beforehand on Strava and I'm good with bearings, so I plan it before I look at the distance and elevation, I then have it on my phone to check but I'm not wearing a watch’ (P14).* |
